# Phenotypic and genomic analysis of a large-scale *Corynebacterium diphtheriae* outbreak among migrant populations in Europe

**DOI:** 10.1101/2023.11.10.23297228

**Authors:** Andreas Hoefer, Helena Seth-Smith, Federica Palma, Stefanie Schindler, Luca Freschi, Alexandra Dangel, Anja Berger, Joshua D’Aeth, Alexander Indra, Norman K. Fry, Daniel Palm, Andreas Sing, Sylvain Brisse, Adrian Egli, the 2022 European diphtheria Consortium

## Abstract

**Background:** Increased numbers of cases of *Corynebacterium diphtheriae* infections were diagnosed in migrant-related facilities of Europe since summer 2022. Most cases involved cutaneous diphtheria, although some respiratory cases and fatalities were reported. A pan-European consortium was created to assess the clinical, epidemiological and microbiological features of this outbreak.

**Methods:** All 363 toxigenic *C. diphtheriae* infection cases from ten European countries were included. Data from case interviews regarding countries of origin and transit routes of migrants were collected. Bacterial isolates underwent whole genome sequencing and antibiotic susceptibility testing. Phylogenetic relationships of outbreak isolates and their antimicrobial resistance genes were studied.

**Results:** Four major genomic clusters were identified, revealing the multiclonal nature of the outbreak. Genes ermX, coding for erythromycin resistance, and genes pbp2m and blaOXA-2 for beta-lactam resistance, were detected in a subset of isolates. Isolates harboring ermX were resistant to erythromycin, and isolates carrying pbp2m were resistant to penicillin, but susceptible to amoxicillin, whereas those carrying blaOXA-2 remained susceptible to beta-lactams. Genomic variation within the four genomic clusters led to estimate their most recent common ancestors between 2017 and 2020.

**Conclusions:** The multi-country distribution of each cluster demonstrated repeated cross-border spread. The increased number of *C. diphtheriae* cases among migrants is a cause for concern, particularly considering antimicrobial resistance phenotypes that threaten the efficacy of first-line treatments. This work provides important knowledge on modern *C. diphtheriae* infections, useful for addressing the reemergence of diphtheria in vulnerable populations and to guide clinical management and measures to control further dissemination.

## Main text

### Resurgence of diphtheria in Europe, 2022

#### Disease background

Diphtheria is a potentially life-threatening and highly transmissible disease, classically causing respiratory illness, but also cutaneous lesions^1^. The main causative agents are toxigenic strains of the bacterium *Corynebacterium diphtheriae*, which express diphtheria toxin (DT), a potent exotoxin that inhibits protein synthesis in eukaryotic cells^2,3^. The resulting tissue damage and systemic toxemia contribute to the disease’s clinical presentation. Diphtheria can manifest in various forms, depending on the site of infection. Respiratory diphtheria is historically the most frequently detected form, presenting with symptoms such as sore throat, low-grade fever, and the formation of pseudo-membranes on the tonsils. Cutaneous diphtheria, characterized by skin ulcers sometimes surrounded by a grayish membrane, is less severe but equally significant for transmission. The production of DT can lead to complications including myocarditis, polyneuropathy, acute kidney disease, and respiratory failure. Non-vaccinated children under five years of age are particularly susceptible. Case fatality rates for respiratory diphtheria can reach 20-40% depending on the affected population, access to medical care and socio-economic conditions. Diphtheria has afflicted human populations since ancient times^4,5^, and devastating epidemics occurred in Europe and North America during the 19th and early 20th centuries, particularly in crowded and unsanitary conditions^6^.

#### Vaccination and disease control

The diphtheria toxoid vaccine has been administered as part of routine childhood immunization since the second World War and was included in the expanded program on immunization (EPI) of the World Health Organization (WHO) in the 1970s. Due to widespread immunization, the incidence of diphtheria in Europe is now very low^7^. Between 2006 and 2021 a mean of 27 cases of *C. diphtheriae* were reported to ECDC within the EU/EEA annually (**Figure S1**). However, diphtheria remains endemic or can become epidemic in regions of the world where vaccine coverage is suboptimal^8,9^. Diagnosed cases in the EU/EEA are often observed among travellers and migrants from such regions^10^.

#### Resurgence

An unusual increase in the number of toxigenic *C. diphtheriae* infections was noted in several European countries from summer 2022 (**Figure S1**)^11–14^. The ECDC published a rapid risk assessment document in October 2022^15^. Here we gathered a consortium of reference laboratories from 10 European countries and investigated the temporal and geographical dynamics and potential source(s) of the outbreak. Using microbiologically confirmed cases, we also assessed the genomic diversity and antimicrobial resistance (AMR) susceptibility of 363 *C. diphtheriae* diphtheria toxin gene *tox*-positive isolates (as defined by PCR).

### Diphtheria in migrant centres in 2022

#### Inclusion criteria and case definition

Cases were retrospectively included in this study for infections with *tox*-gene positive *C. diphtheriae* isolates collected and cultured in ten affected countries, from 1 January to 30 November 2022 (**Figure 1, panel A; Supplementary Appendix**). Whereas the EU/EEA case definition for diphtheria requires both clinically compatible symptoms and in-vitro phenotypic toxigenicity, this study considered individuals harboring *tox* gene positive *C. diphtheriae* as cases, following WHO recommendation for outbreak management of diphtheria and definitions in several EU countries^16,17^ (details in Supplementary appendix). During the study period, we identified 363 isolates of *tox*-gene-bearing *C. diphtheriae* from 362 patients, with the following geographical distribution (**Figure 1, panel B**): Germany (118), Austria (66), UK (59), Switzerland (52), France (30), Belgium (21), Norway (8), The Netherlands (5), Italy (3), and Spain (1).

**Figure 1.**
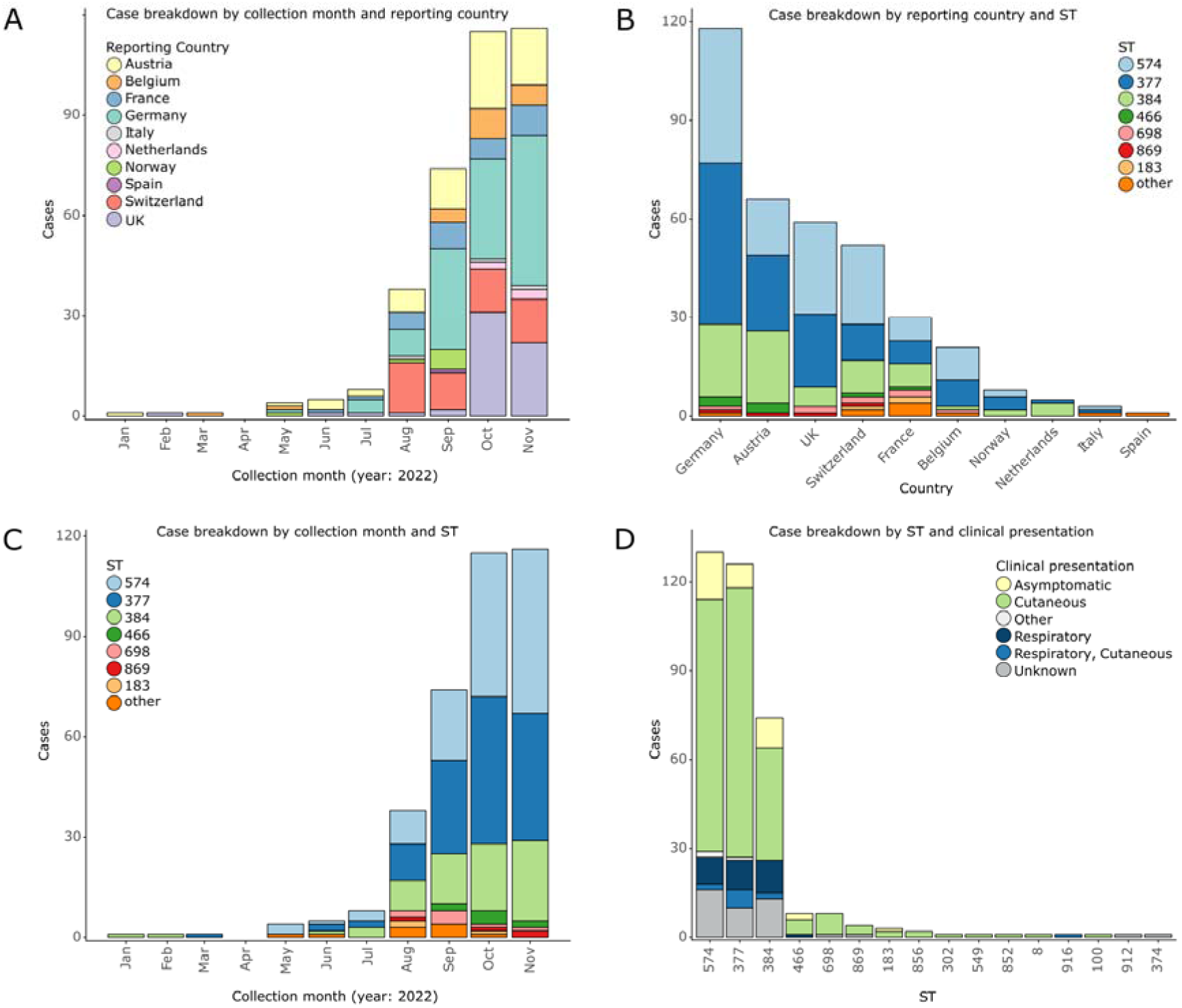
Timeline and geographical distribution of sequence types. **A**, Number of cases detected by reporting countries over the study period; **B**, Number of cases per sequence type, for each reporting country; **C**, Number of cases per sequence types detected over the course of the study period; **D**, Clinical presentation of the 325 cases for which a clinical presentation was reported, for each Sequence Type (STs).

#### Study population

355 (98%) patients were males. Cases had a median age of 18 years at the time of data collection, with 176 (48%) aged between 16 and 20 years. Most of the cases, 348 (96%), had a recent migration history, and/or close contact with migrant populations and/or a recent date of entry into the reporting country, with 174 (48%) cases reported as residents in a migrant center. Once awareness was raised regarding the potential outbreak in 2022, most samples were taken within five days of arrival in the reporting country, in particular as a consequence of enhanced screening activities.

#### Epidemiology of cases and clinical features

From 1 January 2022 to 31 July 2022, a total of 20 cases were reported across six countries. After July, there was a steep increase in cases, with 38 additional cases reported in August, 74 cases in September followed by 115 cases reported in October; prior to plateauing at 116 cases reported by a total of ten countries in November 2022 (**Figure 1, panel A**). The number of toxigenic *C. diphtheriae* cases reported in 2022 marked a drastic increase in cases compared to the average of 27 cases per year from 2006 to 2021 reported to ECDC (**Figure S1**).

Multilocus sequence typing identified 16 Sequence Types (STs), with 330 (91%) of the cases being infected by one of three unrelated sequence types: ST377, ST384 and ST574; these genotypes presented similarly across countries of origin (**see Figure 3**), reporting countries (**Figure 1, panel B**) and over time (**Figure 1, panel C**).

Based on the 325 (90%) cases where clinical presentation was available, 239 (65%) were cutaneous, 43 (12%) were respiratory, including 11 (3.4%) with a pseudo-membrane, and 12 (3.7%) cases were both respiratory and cutaneous. There was no association of clinical symptoms to specific STs (**Figure 1, panel D**). One individual was co-infected by two isolates, a cutaneous ST377 and a respiratory ST384. One respiratory case with a pseudo-membrane had a fatal outcome. We also identified three cases of genital presentation. Owing to the incomplete medical documentation of the patients, it was often difficult to ascertain vaccination status reliably.

#### Migration routes and travel history

During the outbreak investigation, information was collected from case interviews, including country of origin, transit countries, and date of arrival in the reporting country (**Figure 2 and Figure S2**). Out of the 266 (73%) cases where country of origin was available, 19 countries were reported, with 222 (83%) of the cases reported to originate from Afghanistan or Syria. Additionally, a total of 28 transit countries were reported. While there were several cases from Africa and Eastern Europe, most cases followed a Western Balkans migration route (**Figure S2 panel B**).

**Figure 2.**
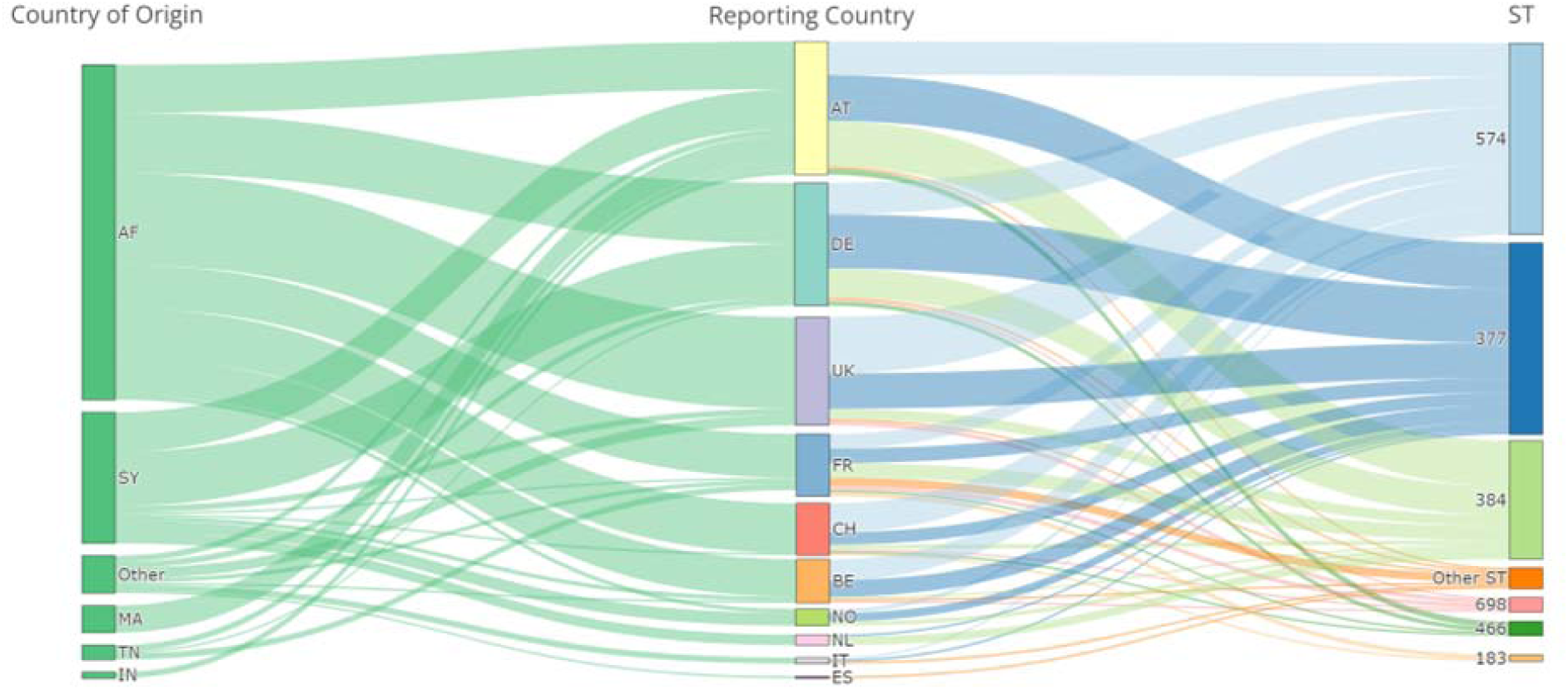
Country of origin *versus* country of reporting and Sequence Types (STs). Alluvial diagram demonstrating the frequency of migration from various countries of origin (column 1) to the reporting countries (column 2) and which sequence types were identified (column 3). AF, Afghanistan; SY, Syria; MA, Morocco; TN, Tunisia; IN, India; AT, Austria; DE, Germany; UK, United Kingdom; FR, France; CH, Switzerland; BE, Belgium; NO, Norway; NL, Netherlands; IT, Italy; and ES, Spain.

### Phenotypic and genomic features of *C. diphtheriae tox*-positive isolates

#### Toxigenicity of isolates

As per our inclusion criteria, the 363 isolates were *tox* gene positive as defined by PCR and confirmed from whole genome sequences. Elek’s test was performed on 306 (84%) isolates, all of which were confirmed to produce the diphtheria toxin. Hence, no non-toxigenic *tox*-gene-bearing (NTTB) isolates were identified over the course of this study^18^, perhaps owing to a lack of selective pressure to interrupt the production of diphtheria toxin, as the migrant population affected was likely largely unvaccinated.

#### Antimicrobial Susceptibility Testing (AST)

Penicillin and erythromycin susceptibility testing was performed on 287 and 282 isolates, respectively, while other antibiotics were tested on subsets of isolates according to country-specific guidelines (**Table S3**). Interpretation was based on the recently established European Committee on Antimicrobial Susceptibility Testing (EUCAST) v13.0 (Jan 2023) clinical breakpoints^19^. For penicillin, 286 (99.7%) of the isolates were susceptible with increased exposure, and for erythromycin, 264 (93.6%) isolates were susceptible. One isolate (0.3%; ID f82239e6, ST183) was resistant to penicillin and meropenem, but not to erythromycin. All 114 isolates that were phenotypically tested against amoxicillin were susceptible. Erythromycin-resistant isolates were in most cases also resistant to ciprofloxacin, tetracycline, doxycycline, and trimethoprim-sulfamethoxazole. Simultaneous phenotypic resistance to both beta-lactams and macrolides was not observed for any isolate. The rates of resistance observed for the other antimicrobial agents (**Table S3**) was high for trimethoprim-sulfamethoxazole (81.1%), tetracycline (32.8%), and ciprofloxacin (22.4%).

#### Genomic diversity of isolates

The nucleotide sequences of 1305 shared core genes were used to generate a phylogenetic tree (**Figure 3**). Ten sublineages (SL, defined as groups of cgMLST profiles with a threshold of 500 allelic mismatches) were discerned, three of which were dominant numerically: SL377 (corresponding to ST377), SL384 (ST384) and SL698 (ST698 and ST574). Within these, four major genomic clusters (GC, defined as groups of cgMLST profiles with a threshold of 25 allelic mismatches) containing >15 genomes were identified: GC795 (n=131) within SL698, GC217 (n=74) corresponding to SL384, and two GCs within SL377: GC817 (n=110) and GC671 (n=17); these two latter GCs were distinct at 65 cgMLST loci. The GCs of the study isolates were newly defined and distinct from previously sequenced isolates. Each of the dominant STs, SLs and GCs were reported from multiple countries (**Figure 3**).

**Figure 3.**
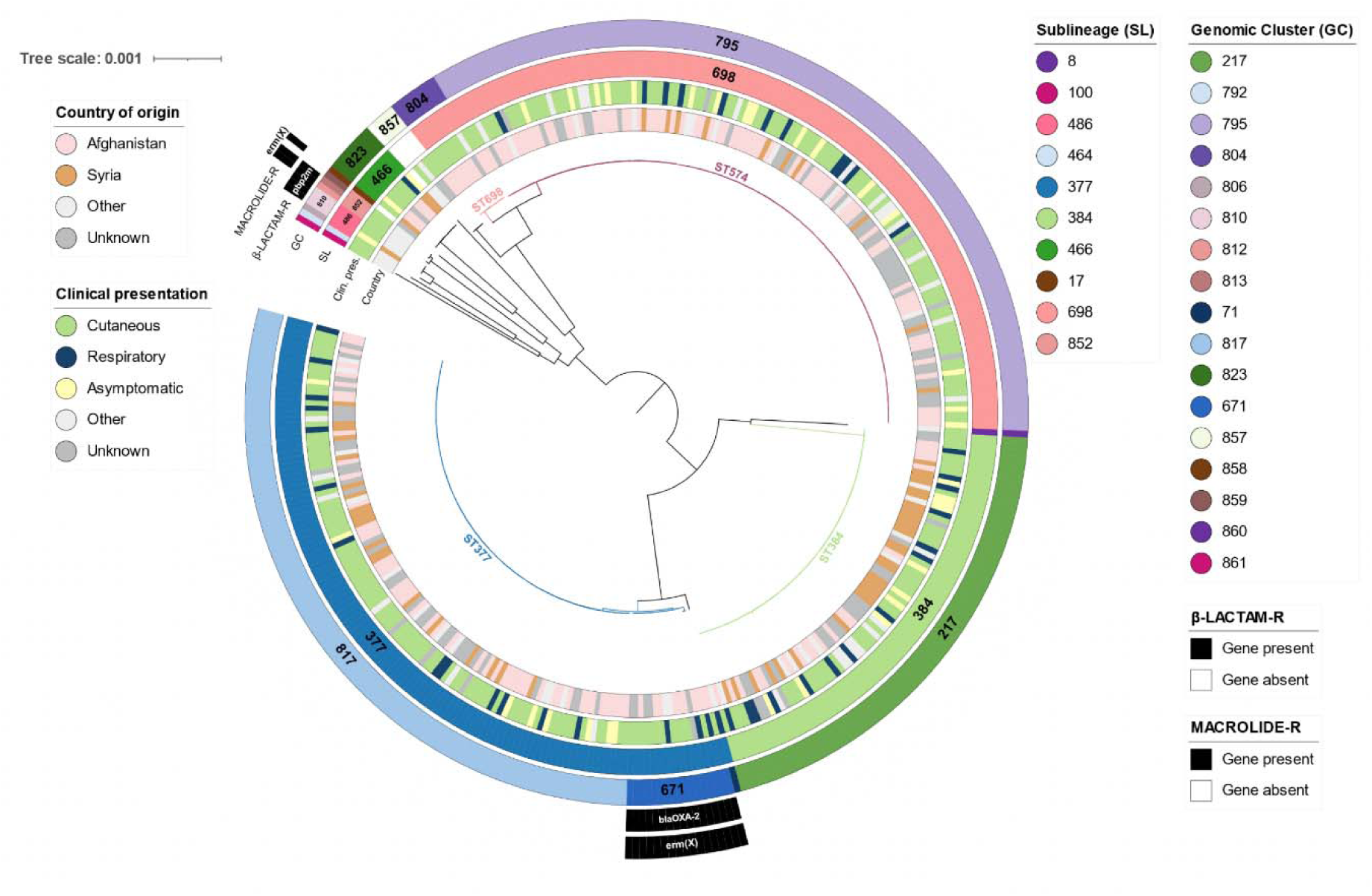
Phylogenetic diversity of the 363 *C. diphtheriae* study isolates. The phylogeny was obtained using core gene sequences and is displayed radially. The branches corresponding to the four major STs are colored. Surrounding metadata circles shown, from inside to outside: country of origin, clinical manifestation, sublineage (SL), genomic cluster (GC), and presence of genes *blaOXA-2*, *pbp2m* potentially conferring resistance to penicillin and gene *ermX* for macrolide resistance (see key).

To put these isolates into wider context, public sequences belonging to SL377, SL384 and SL698 were retrieved from public sequence databases and compared based on cgMLST (**Figure 4**). The most closely related public sequences to SL384 were from the Yemen outbreak^20^; however, they differ from the study isolates by at least 37 cgMLST alleles, and therefore do not belong to the same genomic cluster. Similarly, the most closely related isolates to the European outbreak SL377 isolates were from India, with over 50 cgMLST alleles in distance. No public isolates were found to belong to SL698. Of note, the predominant clusters GC795, GC217 and GC817 were substructured into several variants, each found in several countries (**Figure 4 panels A, B and C**); clearly suggesting genetic diversification into these variants before their cross-border dissemination.

**Figure 4.**
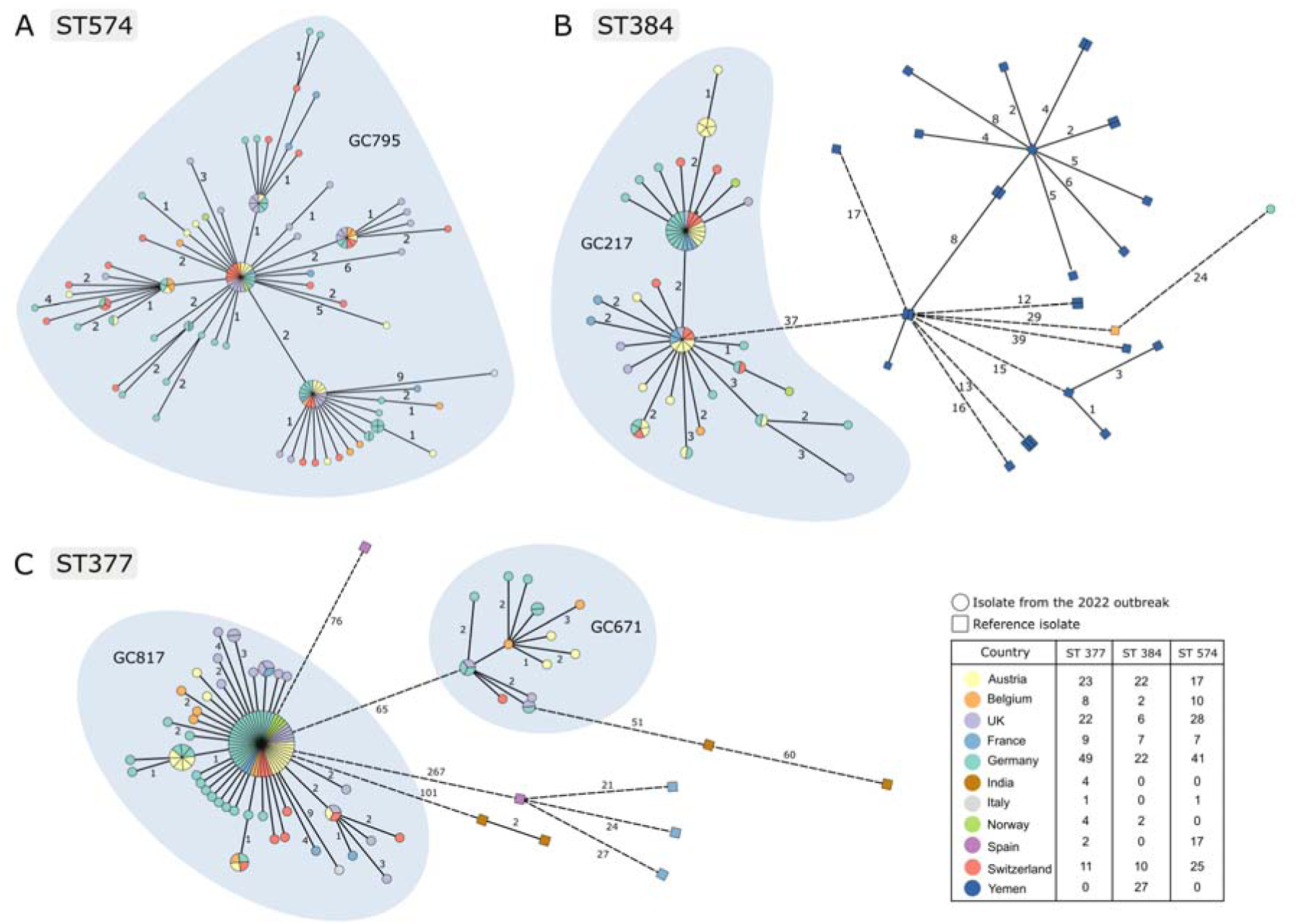
The genetic diversity of the four main genomic clusters. cgMLST based minimum spanning trees of *C. diphtheriae* ST377, ST384 and ST574. Each tree includes the isolates of the outbreak described in this work (circles) as well as reference isolates retrieved from BIGSdb-Pasteur (squares). cgMLST profiles were defined based on the BIGSdb-Pasteur scheme^21^.

#### Detection of antimicrobial resistance genes and integrons

The genome assemblies carried several predicted resistance genes or mutations. In particular *tet(33)*, *dfrA1* and *gyrA* mutations S89F and/or D93Y were observed, and their presence was highly concordant with phenotypic data for tetracycline, trimethoprim and ciprofloxacin, respectively (**Figure S3)**. In addition, *sul1* was found specifically in isolates belonging to SL698 and SL377, and these resisted sulfonamides. A subset of resistant isolates (GC671) carried an integron containing gene *blaOXA-2* as previously described^12^, located in the proximity of *ermX* also found almost exclusively in the smaller SL377 cluster, GC671 (**Figure 3**). The distal position of *blaOXA-2* in the integron may imply it is not expressed, as the tested isolates remained susceptible to penicillin and amoxicillin, as previously reported^12^. The *pbp2m* gene, which decreases susceptibility to penicillin^22^ and the erythromycin-resistance gene *ermX,* were also found in a handful of diverse sporadic genomes (**Figure 3**). The presence of a second integron was also observed, carrying genes for resistance to trimethoprim and aminoglycosides. Most of the SL377 isolates carry this second integron, suggesting that it was acquired before the split that defines the smaller SL377 cluster GC671. This second integron flanks a genomic region where additional chloramphenicol and aminoglycosides resistance genes are found, representing an important antimicrobial resistance genomic region.

#### Diversity and spread of the four genomic clusters

Genome-wide SNP analysis within the four identified genomic clusters, including previously published genomes from the 2022 Switzerland refugee center outbreak^12^ was performed (**Figure 5**). Whereas each cluster was spread across multiple countries, some small phylogenetic subclusters were observed within unique reporting countries, consistent with the cgMLST analysis (**Figure 4**, see *e.g*., the Austria variant at the tip of the vertical branch of the ST384 MStree graph), possibly indicating local chains of transmission.

**Figure 5.**
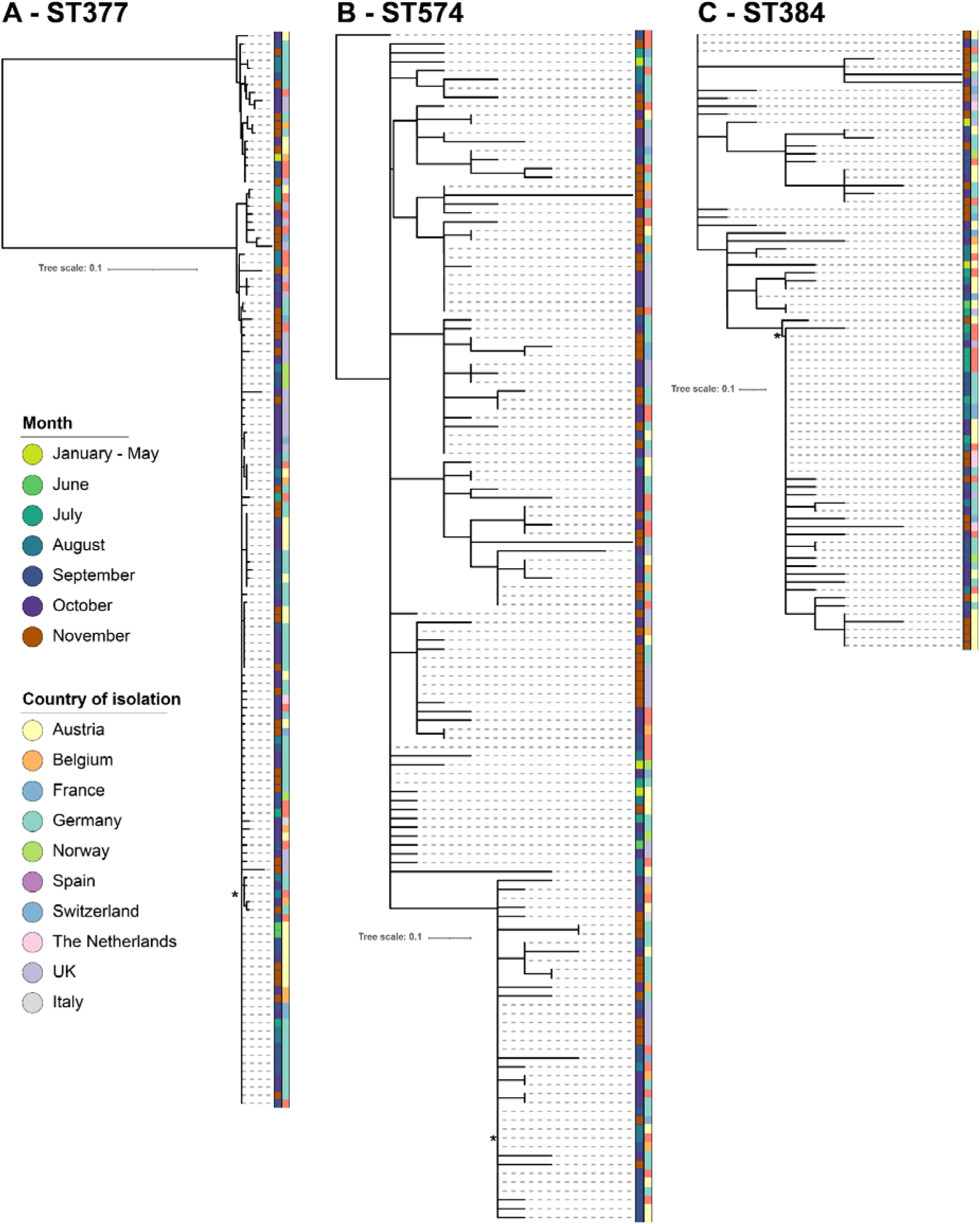
Phylogenies of four main Genomic Clusters (GC), based on whole-genome Single Nucleotide Polymorphisms (SNPs). **A.** ST377 isolates (comprising GC817 and GC671 (n=135)), **B.** ST574 (GC795; n=134); **C.** ST384 (GC217; n=79). The isolates with hybrid reference assemblies used to map against in each tree are indicated with * (accession numbers provided in **Table S2**). Metadata columns represent month of isolation and the reporting country. The tree and metadata were displayed using iTOL v6.7.3. Trees are rooted based on relevant outgroups from the dataset.

The maximal root-to-tip distance within each genomic cluster was 17 SNPs, which underlines their very recent diversification for a single ancestor each. The emergence of the three largest clusters was estimated to have occurred between 2017 and 2020 (**Figure S4**), using a substitution rate of 4.16 nucleotide substitutions per genome per year^20^. The population size history of these clusters was analyzed using skygrowth (**Figure S5**), showing in two cases exponential growth in 2022. Population sizes remained low before 2022, consistent with the clusters not having previously been detected.

### Implications of the 2022 European outbreak

The 2022 outbreak represents the largest rise in diphtheria cases seen in Western Europe in the last 70 years. It occurred primarily among displaced populations entering the region, and with no secondary infections documented among the resident populations.

Where transit data were collected, the vast majority of cases reported migration through the Western Balkan region. Several factors may have facilitated *C. diphtheriae* transmission along this route of migration. While many countries have organized migrant reception centers with dedicated healthcare facilities, large unofficial migrant camps without such amenities can also be found near border regions.^23^ COVID-19 pandemic-related travel restrictions in many European countries further increased population densities in migrant centers and camps^24^, contributing to additional strain on hygiene systems and medical services. These environments are conducive to sustained transmission of *C. diphtheriae,* which may not have been noticed until further down the migration route in central or Western Europe, where disease surveillance and diagnostic capacities are better resourced. Furthermore, among the cases with available clinical presentation data, over 65% presented with cutaneous diphtheria, while only 12% had a respiratory presentation. Cutaneous diphtheria is generally considered a milder form of the disease, less likely to lead to systemic infection, and is less often diagnosed, perhaps explaining in part why this outbreak went unnoticed in Western Balkan transit countries.

The macrolide resistance phenotype observed in the genomic cluster GC671 is of particular concern. Macrolide resistance poses a substantial threat to treatment outcomes and transmission by delaying the administration of effective antibiotics and increases the likelihood of systemic complications requiring diphtheria antitoxin (DAT) treatment. The Centers for Disease Control (CDC), the WHO, and guidelines of many European countries recommend either erythromycin or penicillin as first line treatment (https://www.cdc.gov/diphtheria/clinicians.html). Previous studies reported resistance to erythromycin^25,26^, including a recent association with penicillin reduced susceptibility in *C. diphtheriae* isolates from North and Western Africa^27^.

While most cases were managed with antibiotics only, some respiratory cases required DAT treatment, and one case resulted in a fatal outcome^28^. DAT deployment strategies vary at both national and international levels. While DAT is predominately administered to respiratory cases, there are limited national guidelines defining the circumstances under which DAT treatment is advised. In response to this event, several countries have increased their DAT stockpiles. Increasing the global production of DAT, and maintaining a stable, communal stockpile of DAT that can be rapidly deployed in outbreak situations should be an international public health priority.

### Molecular data underpins informed intervention

Timely generation and sharing of genomic data is recommended as an integral part of outbreak responses^7,29^. Sharing sequencing data among reporting countries allowed other affected countries to determine the degree of relatedness in their circulating isolates, and their commonalities in toxin expression and antimicrobial susceptibility. In the 2022 European outbreak, open sharing of data contributed to public health actions and response options, such as the identification and screening for erythromycin resistance, following the detection of this phenotype in the genomic cluster GC671.

Our study extends earlier local observations^11,12,27^, and has enabled a thorough analysis of the event at continental scale. The simultaneous expansion of multiple GCs across several reporting countries is consistent with genomic investigations of a previous outbreak^20^.

The multi-clonality of this outbreak supports the notion that its origin stems from several distinct emergence events of previously unreported strains, whose transmission was enabled by human contacts among unidentified sustained foci of infection on route to Europe. The age of the common ancestors of the main genomic clusters, estimated between 2017 and 2020, suggests that transmission might have gone unnoticed for some time in these settings/routes and, earlier, in the origin countries of migrants, perhaps with low vaccination levels and limited healthcare infrastructure for diagnosis and surveillance of diphtheria. The simultaneous emergence of several strains and Bayesian analysis results showing a population expansion of the clusters in 2022, suggest that a favorable human to human transmission context played a predominant role, as a simultaneous emergence of particularly virulent or transmissible variants of *C. diphtheriae* appears unlikely.

Whether the 2022 European outbreak reflects a worsening epidemiological situation in countries where diphtheria remains endemic, or sustained transmission along the migration route or in European migrant reception centers, remains unclear and difficult to tell apart with the data currently at hand. Data on migration history and vaccination status were notoriously unreliable. Vaccination coverage in the younger age groups was likely to have been negatively impacted by political instabilities in the countries of origin in the past two decades. This was compounded by the poor availability of reliable medical records for migrants and the absence of a standardized international procedure for interviewing migrant patients. Unfortunately, little information was available from Afghanistan or Syria, although an outbreak in neighboring Pakistan was reported^30^.

Defining the international genomic landscape of *C. diphtheriae*, especially in regions where diphtheria remains endemic, would help future investigations on regional and global dissemination of strains. Laboratory capacity-building efforts in these regions are crucial to enable the diagnostic testing needed to inform patient management, and will also enable wider surveillance efforts to evaluate the public health burden of diphtheria and devise intervention strategies.

### Effective countermeasures

In the months following the study period, there was a drastic reduction in the number of cases, with sporadic cases nevertheless being reported until April 2023^31^. The reduction in the number of cases observed in all reporting countries, can be attributed to several timely and effective countermeasures, including interview-based contact tracing and screening to identify secondary cases. Several countries have initiated chemoprophylaxis of contacts of cases, while others opted for chemoprophylaxis or vaccination campaigns of the wider population within and around migrant centers^32,33^. Furthermore, the lack of secondary cases in European citizens was likely a result of a combination of the timely countermeasures, high vaccination coverage in Western Europe and the relative social isolation of migrant populations. Since the end of the study period there have been two additional fatalities reported in migrants and some European countries have started reporting an increased number of *C. diphtheriae* infections in the early summer of 2023^31^.

This study underscores the continuing importance of global diphtheria vaccination campaigns and programmes, which have mitigated the public health burden of diphtheria for 70 years in Europe, and globally since WHO’s EPI programme. Although childhood vaccination coverage in Europe remains stable, waning immunity and low levels of protection in the elderly may create a vulnerability. The WHO recommends diphtheria vaccination for all children worldwide with a complete primary series plus booster doses. A primary series of three doses of diphtheria toxoid-containing vaccine is recommended followed by three booster doses during childhood and adolescence. The need for additional booster dose later in life to ensure life-long protection remains to be determined^34^, yet recent studies have shown a strong decline in seroprotection among the older adults, leaving them potentially vulnerable to diphtheria caused by both *C. diphtheriae* and *C. ulcerans*^35,36^. While secondary transmission to resident populations was not observed in this study, it remains a possibility. As such, in addition to continued support for childhood and booster vaccinations in European populations, ensuring vaccine boosters in school, adolescent and adult age, and favouring vaccine access in regions where diphtheria remains endemic, will be critical for prevention of sustained diphtheria resurgence in Europe and elsewhere.

## Conclusions

The rise of *C. diphtheriae* case numbers observed among migrants in Western Europe in the second half of 2022 was seemingly halted by rapid response measures, and no forward transmission in the European population was observed. The close relatedness within the genomic clusters of *C. diphtheriae* isolates supports recent transmissions, which may likely have taken place on the migrants route and/or within migrant facilities. The distal origin of this event remains to be determined, even though it might be linked to low vaccination coverage in countries with political unrest and disturbed public health systems that motivate migrations to Europe. This study suggests a number of actions that are needed in Europe to reduce the risk of such outbreaks in future, including: i) improving awareness among migrants, their physicians and relevant personnel in their contact; ii) thorough vaccination protocols in migrants and local population and medical/social care personnel (boosters); iii) timely screening of individuals for symptoms, including non-healing cutaneous lesions; iv) rapid diagnosis of symptomatic persons, with laboratory confirmation of cases by national or international reference centers; and v) performing antimicrobial susceptibility testing and genomic approaches to define appropriate antimicrobial treatment and prophylaxis and decipher local and regional dissemination. Strengthening the implementation of immunisation programmes and seeking to achieve high vaccination coverage (both for primary series and booster doses) in a context of equitable access to vaccination for all groups, including those vulnerable or at risk of being marginalized, remains the key intervention for a protection against diphtheria at population level.

## Data Availability

Accession numbers of genomic data are provided in Table S2.

https://bigsdb.pasteur.fr/cgi-bin/bigsdb/bigsdb.pl?db=pubmlst_diphtheria_isolates&page=query&project_list=17&submit=1

## Acknowledgments

From the Institute of Medical Microbiology, University of Zurich, we thank Nora Köhler, Valéria Pires, Stefan Antener, and Daniel Gander. At Institut Pasteur, we thank Annick Carmi, Sylvie Brémont and Virginie Passet for excellent technical assistance in microbiological confirmation of cases and whole genome sequencing. Within Germany we thank Sabine Lohrer, Jasmin Fräßdorf, Marion Lindermayer, Anne Könitzer, Wolfgang Schmidt, Helga Kocak, Juliane Breitenberger, Andrea Seifarth, and Turgut-Cengiz Dedeoglu, as well as the local, sub-national and national public health authorities, the migrant shelters and the medical laboratories involved in primary bacteriological diagnosis for continuous support. For the UK, we thank colleagues in the UKSHA’s Respiratory and Vaccine Preventable Bacteria Reference Unit and Antimicrobial Resistance and Healthcare Associated Infections Reference Unit for their expert characterization of strains. In Austria, we thank Jasmin Bleier, Petra Hasenberger, and Silke Stadlbauer.

## Funding

The sequencing was funded via the following grants. AE unrestricted grant by the University of Zurich; The Swiss Pathogen Surveillance Platform funded via an NRP72 Swiss National Science Foundation grant; The French National Reference Center for diphtheria is supported by Institut Pasteur and Santé publique France; The German National Consiliary Laboratory for Diphtheria was partly supported by the Bavarian State Ministry of the Health & Care and by the German Federal Ministry of Health via the Robert Koch-Institute and its National Reference Laboratories Network (09-47, FKZ 1369-359).

## Conflict of interest

The authors do not have any conflicts of interests relevant for this publication.

## Ethics statements

**France**: Diphtheria is a notifiable disease in France. Phenotypic and genotypic analyses of bacterial isolates were carried out within the framework of the mandate given to the National Reference Center for Corynebacteria of the Diphtheriae Complex by the Ministry of Health (Public Health France). All French bacteriological samples and associated clinical data are collected, coded, shipped, managed and analyzed according to the French National Reference Center protocols that received approval by French supervisory ethics authority (CNIL, n°1474671).

**Switzerland**: In Switzerland, there is a nationwide ethical approval for outbreak investigation via the Swiss Pathogen Surveillance Platform (SPSP, www.spsp.ch; EKNZ 2019-01291).

**Germany**: Analyses were part of the tasks as National reference center legally covered by the German Infectious Diseases Protection Act (IfSG).

**Austria**: In Austria, outbreak investigation is part of the reference laboratory assignment for mandatory notifiable diseases, including diphtheria, by the Ministry of health. Since all Data gained for this study has been anonymized and gained according to Austrian law (Epidemiegesetz §1. Abs.2, §4, §4a Abs.1. and Abs. 5), no ethical committee approval is needed.

**United Kingdom**: The UK Health Security Agency (and its predecessor organization Public Health England), has legal permission, provided by Regulation 3 of The Health Service (Control of Patient Information) Regulations 2002, to process confidential patient information for national surveillance of communicable diseases (http://www.legislation.gov.uk/uksi/2002/1438/regulation/3/made) and, as such, ethics committee approval is not required.

**Netherlands**: The bacterial isolates belong to the medical microbiology laboratory and were obtained as part of routine clinical care. Since diphtheria is a notifiable disease in the Netherlands, additional phenotypic and genotypic analyses on these isolates were carried out to support outbreak management. To ensure privacy, person identifiers were anonymized before release of any type of data. Furthermore, only the patient’s age in years (not birthdate) and a residential region identifier based on the four digits of the zip code only were collected. Only bacterial isolates and not clinical specimens obtained from patients were used for this study. Since no identifiable personal data was collected and data were analyzed and processed anonymously, written or verbal patient consent was not required. According to the Dutch Medical Research Involving Human Subjects Act (WMO), this study was therefore exempt from review by an Institutional Review Board.

**Norway**: Diphtheria is a notifiable disease in Norway and all the human cases are reported by physicians to the Norwegian Surveillance System for Communicable Diseases (MSIS). Ethical approval was not required as the study was initiated within the legal mandate of the Norwegian Institute of Public Health (NIPH) to investigate and report on infectious disease epidemiology.

**Belgium**: All data accessed in the context of the present study were collected as part of the routine data collection for epidemiological surveillance, as stated in the Public Register dated 25/04/1997. In accordance with §9 of the latter authorization, article 6, §1 and article 9, §2, of the General Data Protection Regulation, no written informed consent from the patients is required for the collection and analysis of epidemiological data and treatment success when processing of personal data is necessary for the performance of a task carried out in the public (health) interest.

**Spain**: Diphtheria is a notifiable disease in Spain. Analyzes were carried out within the tasks trusted to the National Center of Microbiology as stated in “Ley General de Sanidad (Law 14/1986, of April 25)” and the statute of the “Instituto de Salud Carlos III (RD 375 /2001, of April 6 and its subsequent reform, RD 1672/2009, of November 6)”.

**Italy**: Diphtheria is a notifiable disease in Italy and all human case are reported to the Sistema di Sorveglianza delle Malattie Infettive (PREMAL, https://www.seremi.it/sites/default/files/DPAScan0000_1466.pdf<u>), the</u> system for infectious disease surveillance and public health activities of the National Health Service. Analyses (diagnostic confirmation of species and toxicity assay by phenotypic and genotypic methods) of the bacterial isolates, sent by hospital laboratories as part of routine clinical care, are part of the tasks as National Reference Laboratory for Diphtheria, legally covered by the Italian regulation. In this study, only bacterial isolates and not clinical specimens obtained from patients were used and all data were received in an anonymized form. Ethical approval or informed consent were not required as the study was conducted within the legal mandate to investigate and report on infectious disease epidemiology.

## Notes

### Competing Interest Statement

The authors have declared no competing interest.

### Author Declarations

France: Diphtheria is a notifiable disease in France. Phenotypic and genotypic analyses of bacterial isolates were carried out within the framework of the mandate given to the National Reference Center for Corynebacteria of the Diphtheriae Complex by the Ministry of Health (Public Health France). All French bacteriological samples and associated clinical data are collected, coded, shipped, managed and analyzed according to the French National Reference Center protocols that received approval by French supervisory ethics authority (CNIL, number 1474671). Switzerland: In Switzerland, there is a nationwide ethical approval for outbreak investigation via the Swiss Pathogen Surveillance Platform (SPSP, www.spsp.ch; EKNZ 2019-01291). Germany: Analyses were part of the tasks as National reference center legally covered by the German Infectious Diseases Protection Act (IfSG). Austria: In Austria, outbreak investigation is part of the reference laboratory assignment for mandatory notifiable diseases, including diphtheria, by the Ministry of health. Since all Data gained for this study has been anonymized and gained according to Austrian law (Epidemiegesetz 1. Abs.2, 4, 4a Abs.1. and Abs. 5), no ethical committee approval is needed. UK: The UK Health Security Agency (and its predecessor organization Public Health England), has legal permission, provided by Regulation 3 of The Health Service (Control of Patient Information) Regulations 2002, to process confidential patient information for national surveillance of communicable diseases (http://www.legislation.gov.uk/uksi/2002/1438/regulation/3/made) and, as such, ethics committee approval is not required. Netherlands: The bacterial isolates belong to the medical microbiology laboratory and were obtained as part of routine clinical care. Since diphtheria is a notifiable disease in the Netherlands, additional phenotypic and genotypic analyses on these isolates were carried out to support outbreak management. To ensure privacy, person identifiers were anonymized before release of any type of data. Furthermore, only the patient age in years (not birthdate) and a residential region identifier based on the four digits of the zip code only were collected. Only bacterial isolates and not clinical specimensobtained from patients were used for this study. Since no identifiable personal data was collected and data were analyzed and processed anonymously, written or verbal patient consent was not required. According to the Dutch Medical Research Involving Human Subjects Act (WMO), this study was therefore exempt from review by an Institutional Review Board. Norway: Diphtheria is a notifiable disease in Norway and all the human cases are reported by physicians to the Norwegian Surveillance System for Communicable Diseases (MSIS). Ethical approval was not required as the study was initiated within the legal mandate of the Norwegian Institute of Public Health (NIPH) to investigate and report on infectious disease epidemiology. Belgium: All data accessed in the context of the present study were collected as part of the routine data collection for epidemiological surveillance, as stated in the Public Register dated 25/04/1997. In accordance with paragraph 9 of the latter authorization, article 6, paragraph 1 and article 9, paragraph 2, of the General Data Protection Regulation, no written informed consent from the patients is required for the collection and analysis of epidemiological data and treatment success when processing of personal data is necessary for the performance of a task carried out in the public (health) interest. Spain: Diphtheria is a notifiable disease in Spain. Analyzes were carried out within the tasks trusted to the National Center of Microbiology as stated in Ley General de Sanidad (Law 14/1986, of April 25) and the statute of the Instituto de Salud Carlos III (RD 375 /2001, of April 6 and its subsequent reform, RD 1672/2009, of November 6). Italy: Diphtheria is a notifiable disease in Italy and all human case are reported to the Sistema di Sorveglianza delle Malattie Infettive (PREMAL, https://www.seremi.it/sites/default/files/DPAScan0000_1466.pdf), the system for infectious disease surveillance and public health activities of the National Health Service. Analyses (diagnostic confirmation of species and toxicity assay by phenotypic and genotypic methods) of the bacterial isolates, sent by hospital laboratories as part of routine clinical care, are part of the tasks as National Reference Laboratory for Diphtheria, legally covered by the Italian regulation. In this study, only bacterial isolates and not clinical specimens obtained from patients were used and all data were received in an anonymized form. Ethical approval or informed consent were not required as the study was conducted within the legal mandate to investigate and report on infectious disease epidemiology.

