## Supplementary appendix for "Phenotypic and genomic analysis of a large-scale *Corynebacterium diphtheriae* outbreak among migrant populations in Europe"

Affiliations are listed at the end of the author list. Authors listed in alphabetical order.

Alfsnes, Kristian 11

Alves de Sousa, Luis 1

Bacci, Sabrina 1

Badell, Edgar 3

Bengs, Katja 12

Blaschitz, Marion 5

Borrell Pique, Jordi 1

Bressan, Michelle 2

Bruderer, Vera 14

Crestani, Chiara 3

Del Grosso, Maria 15

Delgado, Enrique 1

Didelot, Xavier 16

Dollenmaier, Günter 17

Eletu, Seyi 8

Errico, Giulia 15

Flury, Domenica 18

Fonteneau, Laure 19

Funke, Silvia 1

Füszl, Astrid 5

Heger, Florian 5

Hennart, Mélanie 3

Herrera Leon, Laura 20

Hinic, Vladimira 2

Hobmaier, Bernhard 12

Imkamp, Frank 2

Jacquinet, Stephanie 21

Jost, Geraldine 22

Kuster, Stefan P. 18

Liassine, Nadia 22

Lienhard, Reto 23

Lippert, Kathrin 5

Litt, David 8

Mancini, Stefano 2

Maraglino, Francesco 24

Mariman, Rob 25

Martinetti, Glady 26

Martini, Helena 27

Masa-Calles 20

Monaco, Monica 15

N. Seiffert, Salome 17

Nolte, Oliver 2,17

O'Boyle, Shennae 9

Palamara, Anna Teresa 15

Pleininger, Sonja 5

Pranghofer, Sigrid 13

Purushothaman, Srinithi 2

Roloff, Tim 2

Schibli, Adrian 28

Schmid, Tobias 29

Siddu, Andrea 24

Sprenger, Annika 7

Toubiana, Julie 3

van Meijeren, Dimphey 25

Vaux, Sophie 19

1	European Center for Disease Prevention and Control, Solna, Sweden

2	Institute of Medical Microbiology, University of Zurich, Zurich, Switzerland

3	Institut Pasteur, Université Paris Cité, Biodiversity and Epidemiology of Bacterial Pathogens, Paris, France and National Reference Center for Corynebacteria of the diphtheriae complex, Institut Pasteur, Paris, France

5	Austrian Agency for Health and Food Safety, Vienna, Austria

6	NGS-Coreunit, Public Health Microbiology, Bavarian Health and Food Safety Authority (LGL), Oberschleißheim, Germany

7	National Consiliary Laboratory on Diphtheria, Bavarian Health and Food Safety Authority (LGL), Oberschleißheim, Germany

8	Vaccine Preventable Bacteria Section, Respiratory and Vaccine Preventable Bacteria Reference Unit, Specialised Microbiology and Laboratories Directorate, UK Health Security Agency, London, UK

9	Immunisation and Vaccine Preventable Disease Division, UK Health Security Agency, London, UK

10	Ludwig-Maximilians-Universität, Munich, Germany

11	Norwegian Institute of Public Health (NIPH), Norway

12	Dept. of Public Health Microbiology, Bavarian Health and Food Safety Authority (LGL), Oberschleißheim, Germany

13	Bioanalytica Luzern, Switzerland

14	Medica, Switzerland

15	Istituto Superiore di Sanità, Italy

16	School of Life Sciences and Department of Statistics, University of Warwick, United Kingdom

17	Center for Laboratory Medicine, St Gallen, Switzerland

18	Division of Infectious Diseases and Hospital Epidemiology, Cantonal Hospital of St Gallen, St Gallen, Switzerland

19	Santé Publique France, Saint-Maurice, France

20	Instituto de Salud Carlos III, CIBERESP, Spain

21	Sciensano, Brussels, Belgium

22	Dianalabs, Geneva, Switzerland

23	ADMed Microbiologie La Chaux-de-Fonds, Switzerland

24	Ministry of Health, Italy

25	Rijksinstituut voor Volksgezondheid en Milieu, The Netherlands

26	Ente Ospedaliero Cantonale, Bellinzona, Switzerland

27	Vrije Universiteit Brussel (VUB), Universitair Ziekenhuis Brussel, Belgium

28	Stadtspital Zürich, Switzerland

29	Labor Team, Goldach, Switzerland

### 2.  Supplementary text

*Inclusion criteria and case definition*

Isolates were retrospectively selected for this study as being *tox*-gene positive *C. diphtheriae* isolates as defined by PCR, collected in participating countries from 1st January to 30th November 2022. Although the EU/EEA case definition for diphtheria requires clinically compatible symptoms and phenotypic toxigenicity testing, this study will treat individuals harbouring *tox* gene positive *C. diphtheriae* as cases (even if asymptomatic or polymicrobial cutaneous, and irrespective of toxin production Elek test results).

*Study Population*

While the majority of the cases were among migrants, or had a migrant link, *C. diphtheriae* isolates from the other patients in the participating countries and study period were included to provide a populational context. Among all analysed cases 97% (n=355) were males and 48% (n=176) were aged between 16 and 20 years with a median age of 18 years old at the time of data collection. Of the cases, a country of origin was available for 73% (n=268) of the study population. The majority of the cases (44%, n=160) reported Afghanistan as their country of origin, followed by Syria (18%, n=65) and Morocco (4%, n=13). A total of 19 countries of origin were reported. A breakdown of countries of origin can be found in **Figure S2, Panel A**.

*Sampling methods*

Wound and nasopharyngeal swabs samples were taken from patients and contacts, predominantly asylum seekers in centers across Europe. Date of entry to the reporting country data was collected for 46% (n=169) individuals, of which 72 had their sample taken within five days of arrival, with a median of six days.

*Culturing and Antimicrobial susceptibility testing (AST)*

Microbiological procedures were carried out in accordance with the WHO manual for the laboratory diagnosis of diphtheria^1^. For 79% of the isolates (n=290), phenotypic antimicrobial susceptibility testing (AST) was performed following the EUCAST guidelines and categories were assigned using EUCAST 13.0 breakpoints^2^.

*Whole genome sequencing and bioinformatic analysis*

Isolates underwent whole genome sequencing (WGS), using Illumina (for most isolates) or IonTorrent (for isolates from Italy) platforms. In addition, a subset of isolates were sequenced using ONT technology. A core-gene phylogeny was created based on concatenated alignments of the 1305 core genes^13^ with IQ-Tree 2 (branch lengths corrected to account for recombination using ClonalFrameML v1.12) and represented using iTOL v6.7.3. Analysis by cgMLST was performed using the BIGSdb-Pasteur cgMLST scheme for *C. diphtheriae* (https://bigsdb.pasteur.fr/diphtheria/), which enabled the identification of sublineages (SL) and genomic clusters (GC) with cut-offs of 500 and 25 allele mismatches, respectively. DiphtOscan v1.0.0 (<https://gitlab.pasteur.fr/BEBP/diphtoscan>)^14^ identified the *tox* gene encoding diphtheria toxin, integrons, antimicrobial resistance (AMR) determinants (acquired genes and SNPs) and their genomic context. Comparator reference genomes were also taken from this database. Within-GC analysis was performed using unicycler v0.4.8^15^ hybrid assembled references of Illumina and ONT reads of isolates from within each genomic cluster (see table S2). CLC Genomics Workbench v 22.0.2 was used to perform SNP analysis using parameters that differed from the default as follows: variant calling with 10x minimum coverage, 10 minimum count and 70% minimum frequency, and neighbour joining SNP tree used 10x minimum coverage, 10% minimum coverage, 0 prune distance and including multi-nucleotide variants (MNVs). Tree visualization used iTOL v6.7.3. Data from the Bern (Switzerland) outbreak published in Kofler et al^16^ were also included in the SNP trees of clusters, where appropriate.

Multiple sequence alignments were also generated in CLC Whole Genome Alignment module, exported, and corrected to account for recombination using ClonalFrameML v1.12. Dated phylogenies were computed using BactDating v1.1^17^ under the additive relaxed clock (ARC) model with a mean clock rate of 4.16 substitutions per year as previously estimated^18^. A million iterations were performed, with the first half discarded as burn-in. Demographic inference was performed using skygrowth v0.3.1^19^.

*Other microbiological methods*

For a full breakdown of methods applied in the various affected countries, please see **Table S1**.

*Sequence data availability*

Raw data were made available at the short reads archive of the National Center of Biotechnology Information (NCBI). Accession numbers are provided in **Table S2**. In addition, the genomic assemblies, alleles and profiles and source data of the isolates are available in the project 17 of the BIGSdb-Pasteur diphtheria database at: <https://bigsdb.pasteur.fr/cgi-bin/bigsdb/bigsdb.pl?db=pubmlst_diphtheria_isolates&page=query&project_list=17&submit=1>.

### 3.  Supplementary Tables

##### Table S1. Summary of laboratory and sequencing approaches by country

| **Analysis** | | **Country / institution** |
| --- | --- | --- |
| Culture approach | Tellurite Blood Agar (Hoyle) | Austria, Netherlands, Switzerland (ZLM St. Gall), Spain, UK |
|  | L-Cystine/Tellurite Agar (Tinsdale) | France, UK |
|  | Cystine Tellurite Blood Agar | Belgium, Switzerland (IMM Zurich) |
|  | Serum Tellurite Agar | Germany |
|  | Columbia Blood Agar | Austria, Germany, Italy, Netherlands, Norway, Spain |
|  | MALDI-TOF | Austria, Germany, Italy, UK |
|  | Biochemical tests (e.g. API® CORYNE, BioMerieux) | Austria, Germany, Norway, UK |
|  | Species-specific qPCR | Austria, UK |
| Culture reference | WHO laboratory manual for the diagnosis of diphtheria and other related infections^1^ | Austria, Belgium, France, Germany, Italy, Netherlands, Spain, UK |
|  | In house protocols | Norway, Switzerland (IMM Zurich) |
|  | Manufacturer's instructions | Switzerland (ZLG St. Gall) |
|  | qPCR^3^ | Austria, UK |
| DT-PCR | Yes | Austria, Belgium, France, Germany, Italy, Netherlands, Norway, Spain, Switzerland (IMM Zurich), Switzerland (ZLG St. Gall), UK |
|  | No | / |
| DT-PCR reference | ^4^ | France, Netherlands, Switzerland (ZLG St. Gall) |
|  | ^5^ | Austria |
|  | ^6^ | Germany |
|  | ^7^ | Italy |
|  | ^8^ | Spain |
|  | ^3^ | Norway |
|  | ^9^ (modified) | Switzerland (IMM Zurich) |
|  | In-house primers based on ^10^ | Belgium |
| Elek-test used | Modified Elek-Test | Austria, Belgium, France, Germany, Italy, Spain, Netherlands, Norway, UK |
|  | Optimized Elek-Test | Germany |
|  | Not performed | Switzerland (IMM Zurich), Switzerland (ZLG St. Gall) |
| Elek-test reference | ^11^ (WHO laboratory manual for the diagnosis of diphtheria and other related infections. World Health Organization, 2021.) | Austria, Belgium, France, Germany, Italy, Netherlands, Norway, Spain, UK |
|  | ^12^ (optimized Elek-Test) | Germany |
|  | Not applicable | Switzerland (IMM Zurich), Switzerland (ZLG St. Gall) |
| AMR MIC | Gradient strip tests (ETEST®, biomerieux) | Austria, Belgium, France, Germany, Spain, Switzerland (IMM Zurich) |
|  | Gradient strip tests (Liofilchem®, MIC Test Strips ) | Belgium, Italy |
|  | Not uniformly done | Netherlands, UK |
|  | Not done | Norway, Switzerland (ZLG St. Gall) |
| AMR MIC reference | EUCAST 2023, v 13.0^2^ | Austria, Belgium, France, Germany, Italy, Spain, Netherlands, Switzerland (IMM Zurich), UK |
|  | Not applicable | Norway, Switzerland (ZLG St. Gall) |
| AMR inhibition zone | Disc diffusion test | France, Germany, Switzerland (ZLG St. Gall) |
|  | Not done | Austria, Belgium, Italy, Norway, Spain, Switzerland (IMM Zurich), UK |
|  | Not uniformly done | Netherlands |
| AMR inhibition zone reference | EUCAST 2023, v 13.0^2^ | France, Germany, Switzerland (ZLG St. Gall) |
|  | CA-SFM 2019 | France |
|  | Not applicable | Austria, Belgium, Italy, Norway, Spain, Switzerland (IMM Zurich), UK |
|  | Not uniformly done | Netherlands |
| DNA Extraction | MagAttract HMW kit (QIAGEN) | Austria |
|  | Maxwell® RSC Cultured Cells DNA Kit (Promega) (or protocol IZS) | Belgium |
|  | MasterPure™ Gram-Positive DNA Purification Kit (LGC Biosearch Technologies) | Italy |
|  | QIAGEN Spin Columns (QIAGEN) | France |
|  | Maxwell® 16 LEV Blood DNA Kit (Promega) | Netherlands, Germany |
|  | NZY Tissue gDNA Isolation kit (NZYTech) | Netherlands |
|  | QIAamp DNA Mini Kit (QIAGEN) | Spain, Netherlands, UK |
|  | MagNA Pure Systeme (Roche) | Norway |
|  | Instagene™ Matrix (Bio-Rad) | Switzerland (IMM Zurich) |
|  | QIAsymphony DSP Virus/Pathogen Kit (QIAGEN) | Switzerland (ZLM St. Gall) |
| DNA Extraction reference | Pre-lysis based on Badell et al. 2019 | Austria, Belgium, France, UK |
|  | Pre-lysis based on Dangel et al. 2018 | Germany |
|  | Manufacturer's instructions | Norway, Switzerland (IMM Zurich) |
|  | In house protocol | Italy, Spain, Switzerland (ZLM St. Gall) |
| WGS library method | Nextera XT DNA Library Preparation Kit (Illumina) | Austria, Belgium, France, Netherlands, Spain, UK |
|  | DNA Prep (M) Tagmentation Kit (Illumina) on Beckman Coulter Biomek i7 | Germany |
|  | DNA Prep (M) Tagmentation Kit (Illumina) on Hamilton NGS STAR platform | Switzerland (ZLM St. Gall) |
|  | NEBNext® Fast DNA Library Prep Set for Ion Torrent® (New England Biolabs) | Italy |
|  | KAPA HyperPrep Kit (Roche) | Norway |
|  | QIAseq DNA Library Kit (QIAGEN) | Switzerland (IMM Zurich) |
| WGS platform | NextSeq 500 System (Illumina) | France |
|  | NextSeq 550 System (Illumina) | Germany, Spain, Netherlands |
|  | NextSeq 1000 System (Illumina) | UK |
|  | NextSeq 2000 System (Illumina) | Austria |
|  | NovaSeq 6000 System (Illumina) | Belgium |
|  | MiSeq System (Illumina) | Norway, Switzerland (IMM Zurich) |
|  | MiniSeq System (Illumina) | Switzerland (ZLM St. Gall) |
|  | Ion GeneStudio S5 System (Thermo Fisher Scientific) | Italy |
| WGS read length | 100 bp | UK |
|  | 150 bp | Austria, Switzerland (IMM Zurich), Switzerland (ZLM St. Gall) |
|  | 400 bp | Italy |
|  | 2 x 150 bp | France, Spain, Netherlands |
|  | 2 x 300 bp | Norway |
|  | 2 x 75 bp / 2 x 150 bp | Germany |
|  | 2 x 250 bp / 2 x 150 bp | Belgium |

##### Table S2: Accession numbers

| **TABLE S2: Accession Numbers** | | | | | |
| --- | --- | --- | --- | --- | --- |
| **IsolateID** | **Reporting Country** | Accession Number | Bioproject | Biosample | Nanopore Accession numbers |
| 950003-22 | Austria | SUB13675872 | PRJNA994835 | SAMN36449442 |  |
| 950009-22 | Austria | SUB13675873 | PRJNA994836 | SAMN36449443 |  |
| 950018-22-WH | Austria | SUB13675874 | PRJNA994837 | SAMN36449444 |  |
| 950019-22-WH | Austria | SUB13675875 | PRJNA994838 | SAMN36449445 |  |
| 950032-22 | Austria | SUB13675876 | PRJNA994839 | SAMN36449446 |  |
| 950037-22-WH | Austria | SUB13675877 | PRJNA994840 | SAMN36449447 |  |
| 950039-22-WH | Austria | SUB13675878 | PRJNA994841 | SAMN36449448 |  |
| 950040-22-WH | Austria | SUB13675879 | PRJNA994842 | SAMN36449449 |  |
| 950041-22 | Austria | SUB13675880 | PRJNA994843 | SAMN36449450 |  |
| 950042-22 | Austria | SUB13675881 | PRJNA994844 | SAMN36449451 |  |
| 950049-22 | Austria | SUB13675882 | PRJNA994845 | SAMN36449452 |  |
| 950052-22-WH2 | Austria | SUB13675883 | PRJNA994846 | SAMN36449453 |  |
| 950053-22-WH2 | Austria | SUB13675884 | PRJNA994847 | SAMN36449454 |  |
| 950054-22 | Austria | SUB13675885 | PRJNA994848 | SAMN36449455 |  |
| 950068-22-WH2 | Austria | SUB13675886 | PRJNA994849 | SAMN36449456 |  |
| 950069-22 | Austria | SUB13675887 | PRJNA994850 | SAMN36449457 |  |
| 950071-22 | Austria | SUB13675888 | PRJNA994851 | SAMN36449458 |  |
| 950083-22-WH2 | Austria | SUB13675889 | PRJNA994852 | SAMN36449459 |  |
| 950094-22 | Austria | SUB13675890 | PRJNA994853 | SAMN36449460 |  |
| 950119-22-WH | Austria | SUB13675891 | PRJNA994854 | SAMN36449461 |  |
| 950121-22 | Austria | SUB13675892 | PRJNA994855 | SAMN36449462 |  |
| 950132-22 | Austria | SUB13675893 | PRJNA994856 | SAMN36449463 |  |
| 950133-22-WH | Austria | SUB13675894 | PRJNA994857 | SAMN36449464 |  |
| 950134-22 | Austria | SUB13675895 | PRJNA994858 | SAMN36449465 |  |
| 950135-22 | Austria | SUB13675896 | PRJNA994859 | SAMN36449466 |  |
| 950179-22 | Austria | SUB13675897 | PRJNA994860 | SAMN36449467 |  |
| 950180-22 | Austria | SUB13675898 | PRJNA994861 | SAMN36449468 |  |
| 950181-22-WH | Austria | SUB13675899 | PRJNA994862 | SAMN36449469 |  |
| 950186-22 | Austria | SUB13675900 | PRJNA994863 | SAMN36449470 |  |
| 950187-22-WH | Austria | SUB13675901 | PRJNA994864 | SAMN36449471 |  |
| 950188-22 | Austria | SUB13675902 | PRJNA994865 | SAMN36449472 |  |
| 950189-22 | Austria | SUB13675903 | PRJNA994866 | SAMN36449473 |  |
| 950190-22 | Austria | SUB13675904 | PRJNA994867 | SAMN36449474 |  |
| 950193-22 | Austria | SUB13675905 | PRJNA994868 | SAMN36449475 |  |
| 950199-22 | Austria | SUB13675906 | PRJNA994869 | SAMN36449476 |  |
| 950200-22 | Austria | SUB13675907 | PRJNA994870 | SAMN36449477 |  |
| 950201-22 | Austria | SUB13675908 | PRJNA994871 | SAMN36449478 |  |
| 950205-22 | Austria | SUB13675909 | PRJNA994872 | SAMN36449479 |  |
| 950208-22 | Austria | SUB13675910 | PRJNA994873 | SAMN36449480 |  |
| 950209-22 | Austria | SUB13675911 | PRJNA994874 | SAMN36449481 |  |
| 950212-22 | Austria | SUB13675912 | PRJNA994875 | SAMN36449482 |  |
| 950213-22 | Austria | SUB13675913 | PRJNA994876 | SAMN36449483 |  |
| 950229-22 | Austria | SUB13675914 | PRJNA994877 | SAMN36449484 |  |
| 950232-22 | Austria | SUB13675915 | PRJNA994878 | SAMN36449485 |  |
| 950236-22 | Austria | SUB13675916 | PRJNA994879 | SAMN36449486 |  |
| 950237-22 | Austria | SUB13675917 | PRJNA994880 | SAMN36449487 |  |
| 950239-22 | Austria | SUB13675918 | PRJNA994881 | SAMN36449488 |  |
| 950240-22 | Austria | SUB13675919 | PRJNA994882 | SAMN36449489 |  |
| 950255-22 | Austria | SUB13675920 | PRJNA994883 | SAMN36449490 |  |
| 950260-22 | Austria | SUB13675921 | PRJNA994884 | SAMN36449491 |  |
| 950278-22 | Austria | SUB13675922 | PRJNA994885 | SAMN36449492 |  |
| 950279-1-22-WH2 | Austria | SUB13675923 | PRJNA994886 | SAMN36449493 |  |
| 950280-22 | Austria | SUB13675924 | PRJNA994887 | SAMN36449494 |  |
| 950324-22 | Austria | SUB13675925 | PRJNA994888 | SAMN36449495 |  |
| 950344-22 | Austria | SUB13675926 | PRJNA994889 | SAMN36449496 |  |
| 950346-22-WH | Austria | SUB13675927 | PRJNA994890 | SAMN36449497 |  |
| 950347-17-22 | Austria | SUB13675928 | PRJNA994891 | SAMN36449498 |  |
| 950348-22 | Austria | SUB13675929 | PRJNA994892 | SAMN36449499 |  |
| 950378-WH-22 | Austria | SUB13675930 | PRJNA994893 | SAMN36449500 |  |
| 950474-22 | Austria | SUB13675931 | PRJNA994894 | SAMN36449501 |  |
| 950475-22 | Austria | SUB13675932 | PRJNA994895 | SAMN36449502 |  |
| 950476-22 | Austria | SUB13675933 | PRJNA994896 | SAMN36449503 |  |
| 950483-22 | Austria | SUB13675934 | PRJNA994897 | SAMN36449504 |  |
| 950486-22 | Austria | SUB13675935 | PRJNA994898 | SAMN36449505 |  |
| 950505-22 | Austria | SUB13675936 | PRJNA994899 | SAMN36449506 |  |
| 950542-22 | Austria | SUB13675937 | PRJNA994900 | SAMN36449507 |  |
| DIFT208 | Belgium | SRR25257941 | PRJNA994466 | SAMN36429422 |  |
| DIFT212 | Belgium | SRR25257940 | PRJNA994466 | SAMN36429423 |  |
| DIFT216 | Belgium | SRR25257929 | PRJNA994466 | SAMN36429424 |  |
| DIFT217 | Belgium | SRR25257927 | PRJNA994466 | SAMN36429425 |  |
| DIFT218 | Belgium | SRR25257926 | PRJNA994466 | SAMN36429426 |  |
| DIFT220 | Belgium | SRR25257925 | PRJNA994466 | SAMN36429427 |  |
| DIFT222 | Belgium | SRR25257924 | PRJNA994466 | SAMN36429428 |  |
| DIFT223 | Belgium | SRR25257923 | PRJNA994466 | SAMN36429429 |  |
| DIFT224 | Belgium | SRR25257922 | PRJNA994466 | SAMN36429430 |  |
| DIFT226 | Belgium | SRR25257921 | PRJNA994466 | SAMN36429431 |  |
| DIFT227 | Belgium | SRR25257939 | PRJNA994466 | SAMN36429432 |  |
| DIFT228 | Belgium | SRR25257938 | PRJNA994466 | SAMN36429433 |  |
| DIFT230 | Belgium | SRR25257937 | PRJNA994466 | SAMN36429434 |  |
| DIFT231 | Belgium | SRR25257936 | PRJNA994466 | SAMN36429435 |  |
| DIFT232 | Belgium | SRR25257935 | PRJNA994466 | SAMN36429436 |  |
| DIFT233 | Belgium | SRR25257934 | PRJNA994466 | SAMN36429437 |  |
| DIFT234 | Belgium | SRR25257933 | PRJNA994466 | SAMN36429438 |  |
| DIFT235 | Belgium | SRR25257932 | PRJNA994466 | SAMN36429439 |  |
| DIFT236 | Belgium | SRR25257931 | PRJNA994466 | SAMN36429440 |  |
| DIFT238 | Belgium | SRR25257930 | PRJNA994466 | SAMN36429441 |  |
| DIFT242 | Belgium | SRR25257928 | PRJNA994466 | SAMN36429442 |  |
| FRC1626 | France | ERS14652377 | PRJEB22103 | SAMEA112654733 |  |
| FRC1639 | France | ERS14652385 | PRJEB22103 | SAMEA112654741 |  |
| FRC1662 | France | ERS14652388 | PRJEB22103 | SAMEA112654744 |  |
| FRC1663 | France | ERS14652389 | PRJEB22103 | SAMEA112654745 |  |
| FRC1664 | France | ERS14652390 | PRJEB22103 | SAMEA112654746 |  |
| FRC1670 | France | ERS14652392 | PRJEB22103 | SAMEA112654748 |  |
| FRC1683 | France | ERS14652396 | PRJEB22103 | SAMEA112654752 |  |
| FRC1688 | France | ERS14652398 | PRJEB22103 | SAMEA112654754 |  |
| FRC1701 | France | ERS14652406 | PRJEB22103 | SAMEA112654762 |  |
| FRC1703 | France | ERS14652408 | PRJEB22103 | SAMEA112654764 |  |
| FRC1705 | France | ERS14652410 | PRJEB22103 | SAMEA112654766 |  |
| FRC1716 | France | ERS14652413 | PRJEB22103 | SAMEA112654769 |  |
| FRC1718 | France | ERS14652415 | PRJEB22103 | SAMEA112654771 |  |
| FRC1727 | France | ERS14652417 | PRJEB22103 | SAMEA112654773 |  |
| FRC1728 | France | ERS14652418 | PRJEB22103 | SAMEA112654774 |  |
| FRC1736 | France | ERS14652422 | PRJEB22103 | SAMEA112654778 |  |
| FRC1738 | France | ERS14652423 | PRJEB22103 | SAMEA112654779 |  |
| FRC1746 | France | ERS14652425 | PRJEB22103 | SAMEA112654781 |  |
| FRC1747 | France | ERS14652426 | PRJEB22103 | SAMEA112654782 |  |
| FRC1748 | France | ERS14652427 | PRJEB22103 | SAMEA112654783 |  |
| FRC1750 | France | ERS14652429 | PRJEB22103 | SAMEA112654785 |  |
| FRC1764 | France | ERS14652431 | PRJEB22103 | SAMEA112654787 |  |
| FRC1769 | France | ERS14652433 | PRJEB22103 | SAMEA112654789 |  |
| FRC1770 | France | ERS14652434 | PRJEB22103 | SAMEA112654790 |  |
| FRC1775 | France | ERS14652436 | PRJEB22103 | SAMEA112654792 |  |
| FRC1783 | France | ERS14652438 | PRJEB22103 | SAMEA112654794 |  |
| FRC1784 | France | ERS14652439 | PRJEB22103 | SAMEA112654796 |  |
| FRC1786 | France | ERS14652440 | PRJEB22103 | SAMEA112654797 |  |
| FRC1789 | France | ERS14652442 | PRJEB22103 | SAMEA112654799 |  |
| FRC1791 | France | ERS14652443 | PRJEB22103 | SAMEA112654800 |  |
| KL2070 | Germany | SRR22189703 | PRJNA898270 | SAMN31602461 |  |
| KL2125 | Germany | SRR22189692 | PRJNA898270 | SAMN31602462 |  |
| KL2128 | Germany | SRR22189686 | PRJNA898270 | SAMN31602463 |  |
| KL2129 | Germany | SRR22189685 | PRJNA898270 | SAMN31602464 |  |
| KL2130 | Germany | SRR22189684 | PRJNA898270 | SAMN31602465 |  |
| KL2137 | Germany | SRR22189683 | PRJNA898270 | SAMN31602466 |  |
| KL2139 | Germany | SRR22189682 | PRJNA898270 | SAMN31602467 |  |
| KL2141 | Germany | SRR22189724 | PRJNA898270 | SAMN31602468 |  |
| KL2142 | Germany | SRR22189723 | PRJNA898270 | SAMN31602469 |  |
| KL2143 | Germany | SRR22189722 | PRJNA898270 | SAMN31602470 |  |
| KL2147 | Germany | SRR22189721 | PRJNA898270 | SAMN31602471 |  |
| KL2151 | Germany | SRR22189720 | PRJNA898270 | SAMN31602472 |  |
| KL2158 | Germany | SRR22189719 | PRJNA898270 | SAMN31602473 |  |
| KL2160 | Germany | SRR22189718 | PRJNA898270 | SAMN31602474 |  |
| KL2165 | Germany | SRR22189717 | PRJNA898270 | SAMN31602475 |  |
| KL2168 | Germany | SRR22189716 | PRJNA898270 | SAMN31602476 |  |
| KL2172 | Germany | SRR22189715 | PRJNA898270 | SAMN31602477 |  |
| KL2173 | Germany | SRR25113584 | PRJNA898270 | SAMN36271200 |  |
| KL2176 | Germany | SRR22189713 | PRJNA898270 | SAMN31602478 |  |
| KL2183 | Germany | SRR22189712 | PRJNA898270 | SAMN31602479 |  |
| KL2184 | Germany | SRR22189711 | PRJNA898270 | SAMN31602480 |  |
| KL2187 | Germany | SRR22189710 | PRJNA898270 | SAMN31602481 |  |
| KL2189 | Germany | SRR22189709 | PRJNA898270 | SAMN31602482 |  |
| KL2191 | Germany | SRR25113583 | PRJNA898270 | SAMN36271201 |  |
| KL2192 | Germany | SRR25113572 | PRJNA898270 | SAMN36271202 |  |
| KL2193 | Germany | SRR22189708 | PRJNA898270 | SAMN31602483 |  |
| KL2196 | Germany | SRR22189707 | PRJNA898270 | SAMN31602484 |  |
| KL2197 | Germany | SRR22189706 | PRJNA898270 | SAMN31602485 |  |
| KL2199 | Germany | SRR22189705 | PRJNA898270 | SAMN31602486 |  |
| KL2202 | Germany | SRR22189704 | PRJNA898270 | SAMN31602487 |  |
| KL2203 | Germany | SRR22189702 | PRJNA898270 | SAMN31602488 |  |
| KL2204 | Germany | SRR22189701 | PRJNA898270 | SAMN31602489 |  |
| KL2205 | Germany | SRR25113561 | PRJNA898270 | SAMN36271203 |  |
| KL2206 | Germany | SRR22189700 | PRJNA898270 | SAMN31602490 |  |
| KL2213 | Germany | SRR22189699 | PRJNA898270 | SAMN31602491 |  |
| KL2214 | Germany | SRR22189698 | PRJNA898270 | SAMN31602492 |  |
| KL2219 | Germany | SRR22189697 | PRJNA898270 | SAMN31602493 |  |
| KL2220 | Germany | SRR22189696 | PRJNA898270 | SAMN31602494 |  |
| KL2221 | Germany | SRR22189695 | PRJNA898270 | SAMN31602495 |  |
| KL2224 | Germany | SRR22189694 | PRJNA898270 | SAMN31602496 |  |
| KL2225 | Germany | SRR22189693 | PRJNA898270 | SAMN31602497 |  |
| KL2229 | Germany | SRR25113550 | PRJNA898270 | SAMN36271204 |  |
| KL2230 | Germany | SRR22189691 | PRJNA898270 | SAMN31602498 |  |
| KL2232 | Germany | SRR22189690 | PRJNA898270 | SAMN31602499 |  |
| KL2233 | Germany | SRR22189689 | PRJNA898270 | SAMN31602500 |  |
| KL2234 | Germany | SRR22189688 | PRJNA898270 | SAMN31602501 |  |
| KL2236 | Germany | SRR25113539 | PRJNA898270 | SAMN36271205 |  |
| KL2238 | Germany | SRR25113528 | PRJNA898270 | SAMN36271206 |  |
| KL2239 | Germany | SRR22189687 | PRJNA898270 | SAMN31602502 |  |
| KL2248 | Germany | SRR25113517 | PRJNA898270 | SAMN36271207 |  |
| KL2249 | Germany | SRR25113510 | PRJNA898270 | SAMN36271208 |  |
| KL2252 | Germany | SRR25113509 | PRJNA898270 | SAMN36271209 |  |
| KL2254 | Germany | SRR25113582 | PRJNA898270 | SAMN36271210 |  |
| KL2256 | Germany | SRR25113581 | PRJNA898270 | SAMN36271211 |  |
| KL2257 | Germany | SRR25113580 | PRJNA898270 | SAMN36271212 |  |
| KL2258 | Germany | SRR25113579 | PRJNA898270 | SAMN36271213 |  |
| KL2259 | Germany | SRR25113578 | PRJNA898270 | SAMN36271214 |  |
| KL2260 | Germany | SRR25113577 | PRJNA898270 | SAMN36271215 |  |
| KL2261 | Germany | SRR25113576 | PRJNA898270 | SAMN36271216 |  |
| KL2265 | Germany | SRR25113575 | PRJNA898270 | SAMN36271217 |  |
| KL2266 | Germany | SRR25113574 | PRJNA898270 | SAMN36271218 |  |
| KL2267 | Germany | SRR25113573 | PRJNA898270 | SAMN36271219 |  |
| KL2268 | Germany | SRR25113571 | PRJNA898270 | SAMN36271220 |  |
| KL2270 | Germany | SRR25113570 | PRJNA898270 | SAMN36271221 |  |
| KL2272 | Germany | SRR25113569 | PRJNA898270 | SAMN36271222 |  |
| KL2281 | Germany | SRR25113568 | PRJNA898270 | SAMN36271223 |  |
| KL2282 | Germany | SRR25113567 | PRJNA898270 | SAMN36271224 |  |
| KL2284 | Germany | SRR25113566 | PRJNA898270 | SAMN36271225 |  |
| KL2286 | Germany | SRR25113565 | PRJNA898270 | SAMN36271226 |  |
| KL2289 | Germany | SRR25113564 | PRJNA898270 | SAMN36271227 |  |
| KL2291 | Germany | SRR25113563 | PRJNA898270 | SAMN36271228 |  |
| KL2292 | Germany | SRR25113562 | PRJNA898270 | SAMN36271229 |  |
| KL2293 | Germany | SRR25113560 | PRJNA898270 | SAMN36271230 |  |
| KL2299 | Germany | SRR25113559 | PRJNA898270 | SAMN36271231 |  |
| KL2300 | Germany | SRR25113558 | PRJNA898270 | SAMN36271232 |  |
| KL2301 | Germany | SRR25113557 | PRJNA898270 | SAMN36271233 |  |
| KL2302 | Germany | SRR25113556 | PRJNA898270 | SAMN36271234 |  |
| KL2303 | Germany | SRR25113555 | PRJNA898270 | SAMN36271235 |  |
| KL2304 | Germany | SRR25113554 | PRJNA898270 | SAMN36271236 |  |
| KL2305 | Germany | SRR25113553 | PRJNA898270 | SAMN36271237 |  |
| KL2306 | Germany | SRR25113552 | PRJNA898270 | SAMN36271238 |  |
| KL2308 | Germany | SRR25113551 | PRJNA898270 | SAMN36271239 |  |
| KL2310 | Germany | SRR25113549 | PRJNA898270 | SAMN36271240 |  |
| KL2311 | Germany | SRR25113548 | PRJNA898270 | SAMN36271241 |  |
| KL2312 | Germany | SRR25113547 | PRJNA898270 | SAMN36271242 |  |
| KL2314 | Germany | SRR25113546 | PRJNA898270 | SAMN36271243 |  |
| KL2315 | Germany | SRR25113545 | PRJNA898270 | SAMN36271244 |  |
| KL2319 | Germany | SRR25113544 | PRJNA898270 | SAMN36271245 |  |
| KL2320 | Germany | SRR25113543 | PRJNA898270 | SAMN36271246 |  |
| KL2321 | Germany | SRR25113542 | PRJNA898270 | SAMN36271247 |  |
| KL2322 | Germany | SRR25113541 | PRJNA898270 | SAMN36271248 |  |
| KL2326 | Germany | SRR25113540 | PRJNA898270 | SAMN36271249 |  |
| KL2327 | Germany | SRR25113538 | PRJNA898270 | SAMN36271250 |  |
| KL2328 | Germany | SRR25113537 | PRJNA898270 | SAMN36271251 |  |
| KL2329 | Germany | SRR25113536 | PRJNA898270 | SAMN36271252 |  |
| KL2331 | Germany | SRR25113535 | PRJNA898270 | SAMN36271253 |  |
| KL2333 | Germany | SRR25113534 | PRJNA898270 | SAMN36271254 |  |
| KL2336 | Germany | SRR25113533 | PRJNA898270 | SAMN36271255 |  |
| KL2337 | Germany | SRR25113532 | PRJNA898270 | SAMN36271256 |  |
| KL2338 | Germany | SRR25113531 | PRJNA898270 | SAMN36271257 |  |
| KL2341 | Germany | SRR25113530 | PRJNA898270 | SAMN36271258 |  |
| KL2342 | Germany | SRR25113529 | PRJNA898270 | SAMN36271259 |  |
| KL2344 | Germany | SRR25113527 | PRJNA898270 | SAMN36271260 |  |
| KL2345 | Germany | SRR25113526 | PRJNA898270 | SAMN36271261 |  |
| KL2346 | Germany | SRR25113525 | PRJNA898270 | SAMN36271262 |  |
| KL2348A | Germany | SRR25113524 | PRJNA898270 | SAMN36271263 |  |
| KL2351 | Germany | SRR25113523 | PRJNA898270 | SAMN36271264 |  |
| KL2353 | Germany | SRR25113522 | PRJNA898270 | SAMN36271265 |  |
| KL2354 | Germany | SRR25113521 | PRJNA898270 | SAMN36271266 |  |
| KL2355 | Germany | SRR25113520 | PRJNA898270 | SAMN36271267 |  |
| KL2357 | Germany | SRR25113519 | PRJNA898270 | SAMN36271268 |  |
| KL2358 | Germany | SRR25113518 | PRJNA898270 | SAMN36271269 |  |
| KL2359 | Germany | SRR25113516 | PRJNA898270 | SAMN36271270 |  |
| KL2361 | Germany | SRR25113515 | PRJNA898270 | SAMN36271271 |  |
| KL2362 | Germany | SRR25113514 | PRJNA898270 | SAMN36271272 |  |
| KL2385 | Germany | SRR25113513 | PRJNA898270 | SAMN36271273 |  |
| KL2386 | Germany | SRR25113512 | PRJNA898270 | SAMN36271274 |  |
| KL2387 | Germany | SRR25113511 | PRJNA898270 | SAMN36271275 |  |
| CD197 | Italy | ERS16055683 | PRJEB64352 | SAMEA114072281 |  |
| CD201 | Italy | ERS16055684 | PRJEB64352 | SAMEA114072282 |  |
| CD204 | Italy | ERS16055685 | PRJEB64352 | SAMEA114072283 |  |
| NL2022_A001 | The Netherlands | JAUOOJ000000000 | PRJNA996478 | SAMN36575637 |  |
| NL2022_A002 | The Netherlands | JAUOOL000000000 | PRJNA996478 | SAMN36575638 |  |
| NL2022_A003 | The Netherlands | JAUOOK000000000 | PRJNA996478 | SAMN36575639 |  |
| NL2022_A004 | The Netherlands | JAUOOH000000000 | PRJNA996478 | SAMN36575640 |  |
| NL2022_A005 | The Netherlands | JAUOOI000000000 | PRJNA996478 | SAMN36575641 |  |
| NTD1 | Norway | ERS15960518 | PRJEB63682 |  |  |
| NTD2 | Norway | ERS15960519 | PRJEB63683 |  |  |
| NTD3 | Norway | ERS15960520 | PRJEB63684 |  |  |
| NTD4 | Norway | ERS15960521 | PRJEB63685 |  |  |
| NTD5 | Norway | ERS15960522 | PRJEB63686 |  |  |
| NTD6skin | Norway | ERS15960523 | PRJEB63687 |  |  |
| NTD6throat | Norway | ERS15960524 | PRJEB63688 |  |  |
| NTD7 | Norway | ERS15960525 | PRJEB63689 |  |  |
| Cd20220012 | Spain | SRR26038076 | PRJNA1015574 | SAMN37358071 |  |
| f82239e6 | Switzerland | ERS14359421 | PRJEB57872 | ERR10679278 |  |
| 8678946e | Switzerland | ERS14359424 | PRJEB57872 | ERR10679255 |  |
| e314b4fb | Switzerland | ERS14359425 | PRJEB57872 | ERR10679272 |  |
| 22cf7fbb | Switzerland | ERS14359418 | PRJEB57872 | ERR10679236 |  |
| 9ca47288 | Switzerland | ERS14359422 | PRJEB57872 | ERR10679259 |  |
| b308b3ee | Switzerland | ERS14359420 | PRJEB57872 | ERR10679264 |  |
| b5c7c6d7 | Switzerland | ERS14359423 | PRJEB57872 | ERR10679265 |  |
| 7ed8f9b2 | Switzerland | ERS14359429 | PRJEB57872 | ERR10679254 |  |
| f37f576b | Switzerland | ERS14359451 | PRJEB57872 | ERR10679275 |  |
| f3fd6441 | Switzerland | ERS14359450 | PRJEB57872 | ERR10679277 |  |
| 30140a2e | Switzerland | ERS14359452 | PRJEB57872 | ERR10679239 |  |
| 7cf10d40 | Switzerland | ERS14359458 | PRJEB57872 | ERR10679253 |  |
| df4929a5 | Switzerland | ERS14359457 | PRJEB57872 | ERR10679271 |  |
| 9edc0788 | Switzerland | ERS14314602 | PRJEB57872 | ERR10639949 |  |
| ab7184bc | Switzerland | ERS14359460 | PRJEB57872 | ERR10679263 |  |
| 297dd70a | Switzerland | ERS14359443 | PRJEB57872 | ERR10679238 |  |
| 6a3548aa | Switzerland | ERS14359446 | PRJEB57872 | ERR10679247 |  |
| 3aaa6cf3 | Switzerland | ERS14470156 | PRJEB57872 | ERR10802408 |  |
| 9616317d | Switzerland | ERS14359445 | PRJEB57872 | ERR10679257 |  |
| c48d9d4e | Switzerland | ERS14470161 | PRJEB57872 | ERR10802416 |  |
| dd77a507 | Switzerland | ERS14470162 | PRJEB57872 | ERR10802420 |  |
| b98973d0 | Switzerland | ERS14359436 | PRJEB57872 | ERR10679266 |  |
| 54d550d0 | Switzerland | ERS14359435 | PRJEB57872 | ERR10679245 |  |
| 42606c6f | Switzerland | ERS14470152 | PRJEB57872 | ERR10802409 |  |
| 94c3d8af | Switzerland | ERS14359417 | PRJEB57872 | ERR10679256 |  |
| dbde5ce5 | Switzerland | ERS14314579 | PRJEB57872 | ERR10639956 | Pending |
| c52a6253 | Switzerland | ERS14314580 | PRJEB57872 | ERR10639952 |  |
| f244bdaf | Switzerland | ERS14314578 | PRJEB57872 | ERR10639958 |  |
| a17b6a40 | Switzerland | ERS14314582 | PRJEB57872 | ERR10639950 | Pending |
| c6aa67fa | Switzerland | ERS14314583 | PRJEB57872 | ERR10639953 |  |
| 20e8f080 | Switzerland | ERS14314586 | PRJEB57872 | ERR10639932 | Pending |
| 4a2da6f7 | Switzerland | ERS14314587 | PRJEB57872 | ERR10639939 |  |
| 722d66ab | Switzerland | ERS14314588 | PRJEB57872 | ERR10639942 |  |
| 31b735af | Switzerland | ERS14314596 | PRJEB57872 | ERR10639933 |  |
| f3a4be5f | Switzerland | ERS14359440 | PRJEB57872 | ERR10679276 |  |
| ceaf779c | Switzerland | ERS14470151 | PRJEB57872 | ERR10802418 |  |
| 9bf9deca | Switzerland | ERS14359456 | PRJEB57872 | ERR10679258 |  |
| 7871f74d | Switzerland | ERS14470154 | PRJEB57872 | ERR10802413 |  |
| 7466c630 | Switzerland | ERS14359454 | PRJEB57872 | ERR10679249 |  |
| 08bfa5ab | Switzerland | ERS14314598 | PRJEB57872 | ERR10639931 |  |
| f8bc126e | Switzerland | ERS14359459 | PRJEB57872 | ERR10679279 |  |
| cd4b0c09 | Switzerland | ERS14470153 | PRJEB57872 | ERR10802417 |  |
| 4be88993 | Switzerland | ERS14359462 | PRJEB57872 | ERR10679244 |  |
| 72b93d78 | Switzerland | ERS14359444 | PRJEB57872 | ERR10679248 |  |
| dea539c9 | Switzerland | ERS14470155 | PRJEB57872 | ERR10802422 |  |
| a323ea36 | Switzerland | ERS14359448 | PRJEB57872 | ERR10679261 |  |
| e6dbd65c | Switzerland | ERS14470160 | PRJEB57872 | ERR10802423 |  |
| a859c682 | Switzerland | ERS14470163 | PRJEB57872 | ERR10802414 |  |
| 42c5f5f5 | Switzerland | ERS14470164 | PRJEB57872 | ERR10802410 |  |
| a90aa8d4 | Switzerland | ERS14470157 | PRJEB57872 | ERR10802415 |  |
| 49a3398f | Switzerland | ERS14470159 | PRJEB57872 | ERR10802411 |  |
| d920e53b | Switzerland | ERS14470165 | PRJEB57872 | ERR10802419 |  |
| UK01 | UK | ERR11635567 | PRJEB63627 |  |  |
| UK02 | UK | ERR11635568 | PRJEB63627 |  |  |
| UK03 | UK | ERR11635569 | PRJEB63627 |  |  |
| UK04 | UK | ERR11635571 | PRJEB63627 |  |  |
| UK05 | UK | ERR11635572 | PRJEB63627 |  |  |
| UK06 | UK | ERR11635574 | PRJEB63627 |  |  |
| UK07 | UK | ERR11635575 | PRJEB63627 |  |  |
| UK08 | UK | ERR11635576 | PRJEB63627 |  |  |
| UK09 | UK | ERR11635577 | PRJEB63627 |  |  |
| UK10 | UK | ERR11635578 | PRJEB63627 |  |  |
| UK11 | UK | ERR11635579 | PRJEB63627 |  |  |
| UK12 | UK | ERR11635580 | PRJEB63627 |  |  |
| UK13 | UK | ERR11635581 | PRJEB63627 |  |  |
| UK14 | UK | ERR11635914 | PRJEB63627 |  |  |
| UK15 | UK | ERR11635614 | PRJEB63627 |  |  |
| UK16 | UK | ERR11635616 | PRJEB63627 |  |  |
| UK17 | UK | ERR11635617 | PRJEB63627 |  |  |
| UK18 | UK | ERR11635618 | PRJEB63627 |  |  |
| UK19 | UK | ERR11635619 | PRJEB63627 |  |  |
| UK20 | UK | ERR11635620 | PRJEB63627 |  |  |
| UK21 | UK | ERR11635621 | PRJEB63627 |  |  |
| UK22 | UK | ERR11635624 | PRJEB63627 |  |  |
| UK23 | UK | ERR11635625 | PRJEB63627 |  |  |
| UK24 | UK | ERR11635626 | PRJEB63627 |  |  |
| UK25 | UK | ERR11635696 | PRJEB63627 |  |  |
| UK26 | UK | ERR11635698 | PRJEB63627 |  |  |
| UK27 | UK | ERR11635699 | PRJEB63627 |  |  |
| UK28 | UK | ERR11635700 | PRJEB63627 |  |  |
| UK29 | UK | ERR11635701 | PRJEB63627 |  |  |
| UK30 | UK | ERR11635702 | PRJEB63627 |  |  |
| UK31 | UK | ERR11635703 | PRJEB63627 |  |  |
| UK32 | UK | ERR11635704 | PRJEB63627 |  |  |
| UK33 | UK | ERR11635705 | PRJEB63627 |  |  |
| UK34 | UK | ERR11635706 | PRJEB63627 |  |  |
| UK35 | UK | ERR11635707 | PRJEB63627 |  |  |
| UK36 | UK | ERR11635708 | PRJEB63627 |  |  |
| UK37 | UK | ERR11635709 | PRJEB63627 |  |  |
| UK38 | UK | ERR11635711 | PRJEB63627 |  |  |
| UK39 | UK | ERR11635714 | PRJEB63627 |  |  |
| UK40 | UK | ERR11635717 | PRJEB63627 |  |  |
| UK41 | UK | ERR11635719 | PRJEB63627 |  |  |
| UK42 | UK | ERR11635725 | PRJEB63627 |  |  |
| UK43 | UK | ERR11635728 | PRJEB63627 |  |  |
| UK44 | UK | ERR11635730 | PRJEB63627 |  |  |
| UK45 | UK | ERR11635735 | PRJEB63627 |  |  |
| UK46 | UK | ERR11635741 | PRJEB63627 |  |  |
| UK47 | UK | ERR11635744 | PRJEB63627 |  |  |
| UK48 | UK | ERR11635746 | PRJEB63627 |  |  |
| UK49 | UK | ERR11635752 | PRJEB63627 |  |  |
| UK51 | UK | ERR11635756 | PRJEB63627 |  |  |
| UK52 | UK | ERR11635758 | PRJEB63627 |  |  |
| UK53 | UK | ERR11635766 | PRJEB63627 |  |  |
| UK54 | UK | ERR11635767 | PRJEB63627 |  |  |
| UK55 | UK | ERR11635769 | PRJEB63627 |  |  |
| UK56 | UK | ERR11635774 | PRJEB63627 |  |  |
| UK57 | UK | ERR11635778 | PRJEB63627 |  |  |
| UK58 | UK | ERR11635779 | PRJEB63627 |  |  |
| UK59 | UK | ERR11635780 | PRJEB63627 |  |  |
| UK60 | UK | ERR11635781 | PRJEB63627 |  |  |

##### Table S3. Phenotypic antimicrobial susceptibility testing results

|  | **No. of isolates tested** | **Susceptible** | | **Susceptible increased exp*** | | **Resistant** | |
| --- | --- | --- | --- | --- | --- | --- | --- |
|  |  | **n** | **%** | **n** | **%** | **n** | **%** |
| Penicillin | 287 | 0 | 0.0% | 286 | 99.7% | 1 | 0.3% |
| Erythromycin | 282 | 264 | 93.6% | 0 | 0.0% | 18 | 6.4% |
| Amoxicillin | 114 | 114 | 100.0% | 0 | 0.0% | 0 | 0.0% |
| Meropenem | 61 | 60 | 98.4% | 0 | 0.0% | 1 | 1.6% |
| Tetracycline | 180 | 121 | 67.2% | 0 | 0.0% | 59 | 32.8% |
| Trimethoprim-Sulfamethoxazole | 175 | 33 | 18.9% | 0 | 0.0% | 142 | 81.1% |
| Ciprofloxacin | 255 | 0 | 0.0% | 198 | 77.6% | 57 | 22.4% |
| Linezolid | 181 | 181 | 100.0% | 0 | 0.0% | 0 | 0.0% |
| Clindamycin | 227 | 199 | 87.7% | 0 | 0.0% | 28 | 12.3% |
| Rifampicin | 153 | 153 | 100.0% | 0 | 0.0% | 0 | 0.0% |
| Doxycycline | 72 | 47 | 65.3% | 0 | 0.0% | 25 | 34.7% |
| Cefotaxime | 6 | 0 | 0.0% | 6 | 100.0% | 0 | 0.0% |

**Susceptible at increased exposure*

### 4.  Supplementary Figures

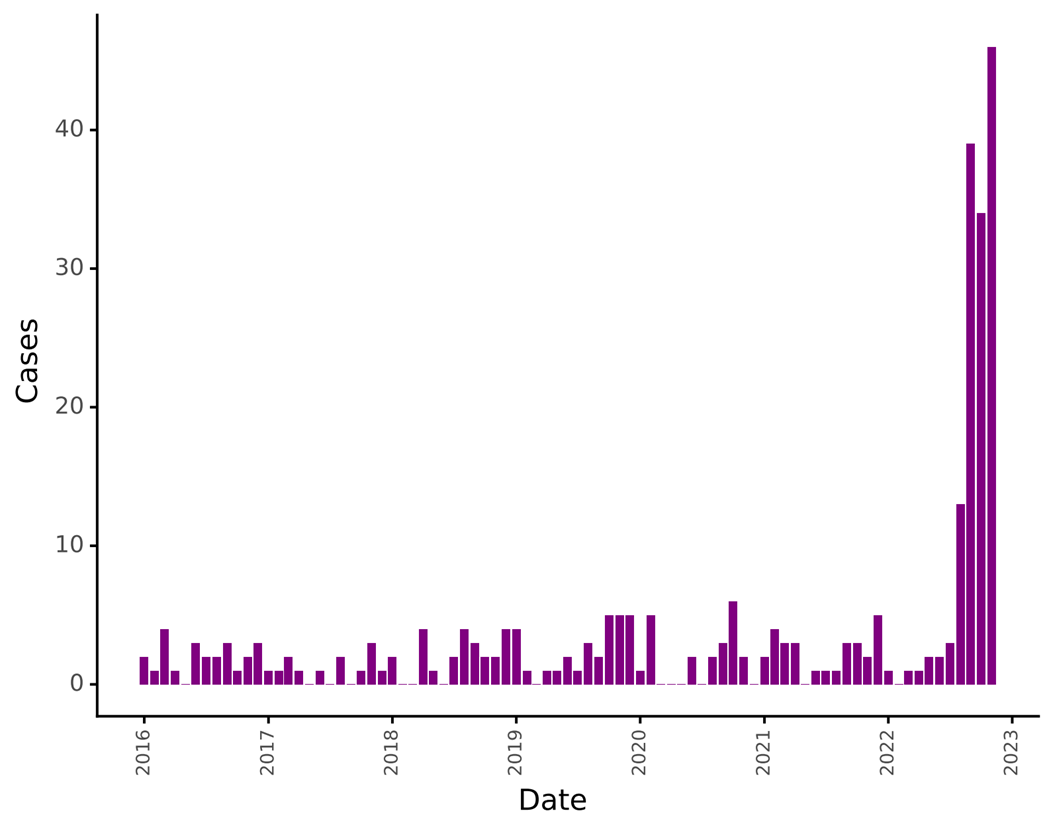

##### **Figure S1. Historical reported cases of diphtheria in EU/EEA.**

Data retrieved from ECDC Atlas on C. diphtheriae cases from 2016 until November 2022. Case counts do not include cases from Switzerland and also may not reflect all cases as collected by the consortium meaning the case count in 2022 is likely to be underestimated here. UK data was only available until 2021.

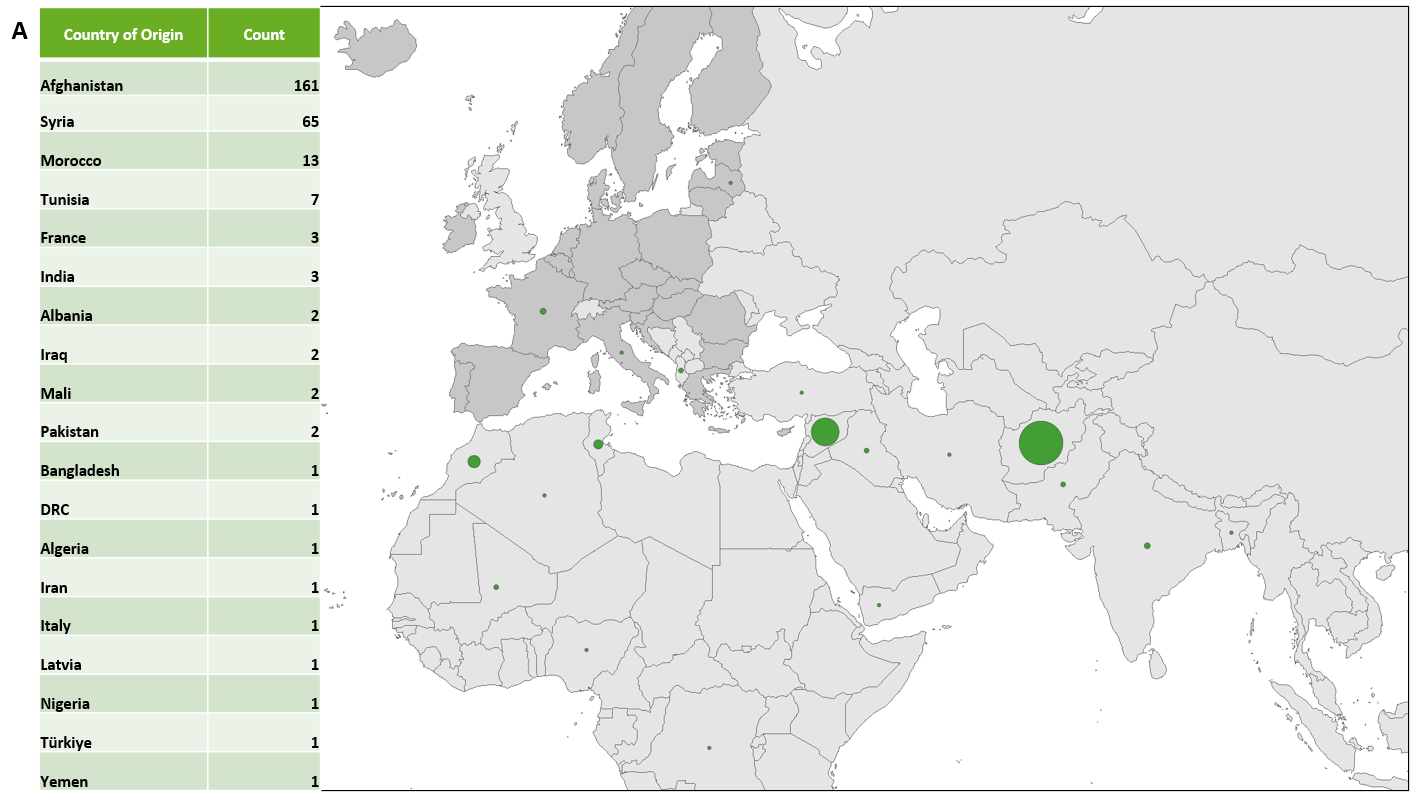

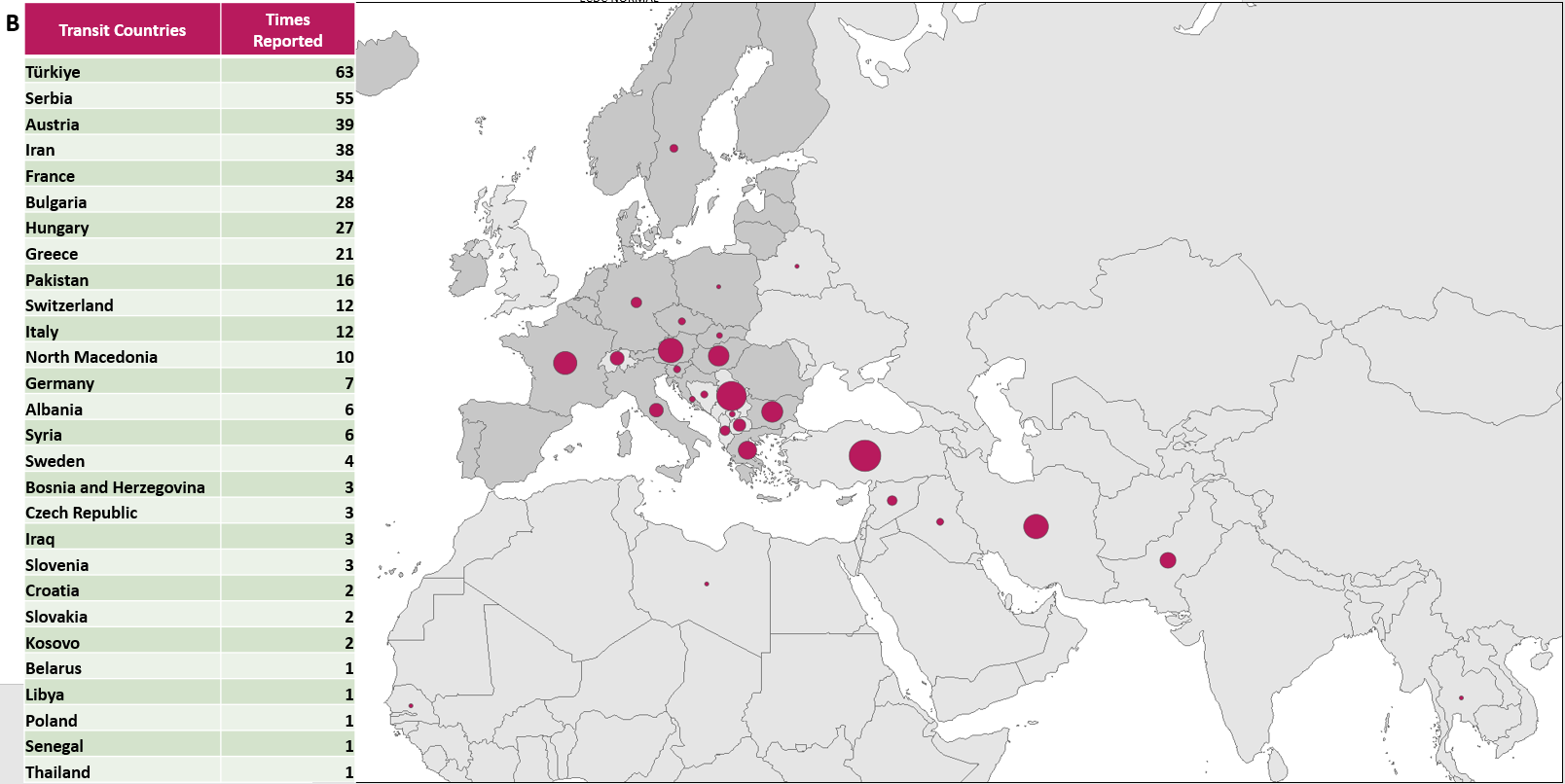

##### **Figure S2: Reported countries of origin and transit countries**.

A. countries of origin as described by cases during interviews by reporting country authorities. B. cumulative (includes multiple countries if reported by a single individual) transit countries described by cases during interviews by reporting country authorities.

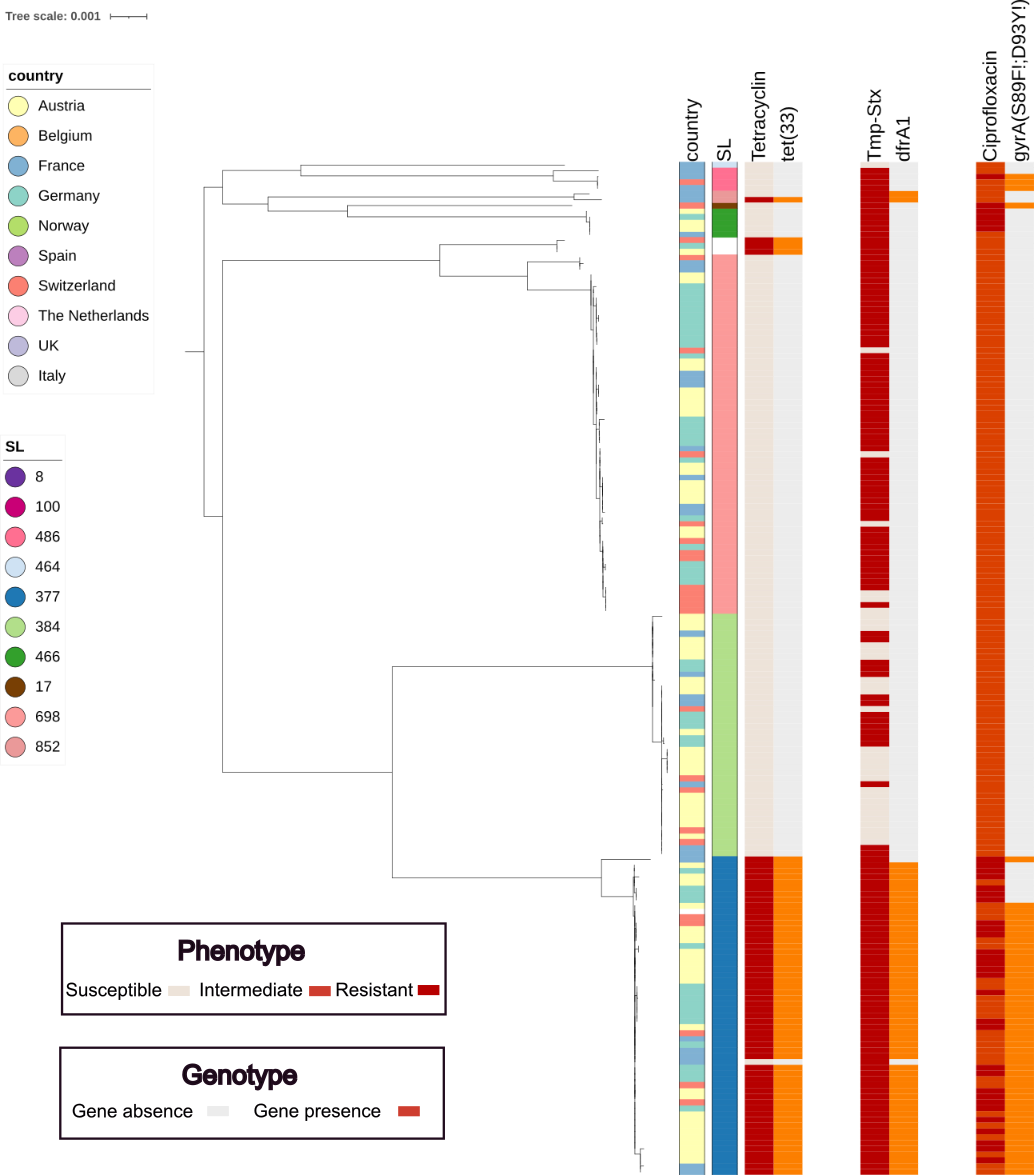

##### **Figure S3. Phylogenetic distribution of resistance profiles.**

Comparison of phenotypic resistance testing with resistance genotype of selected agents. The three last pairs of columns correspond to tetracycline phenotype and tet(33) detection, trimethoprim-sulfamethoxazole phenotype and dfrA1 detection, and ciprofloxacin phenotype and mutations in *gyrA* leading to amino acid changes S89F and/or D93Y in the gyrase subunit A protein. Reporting country and sublineage (SL) are also indicated.

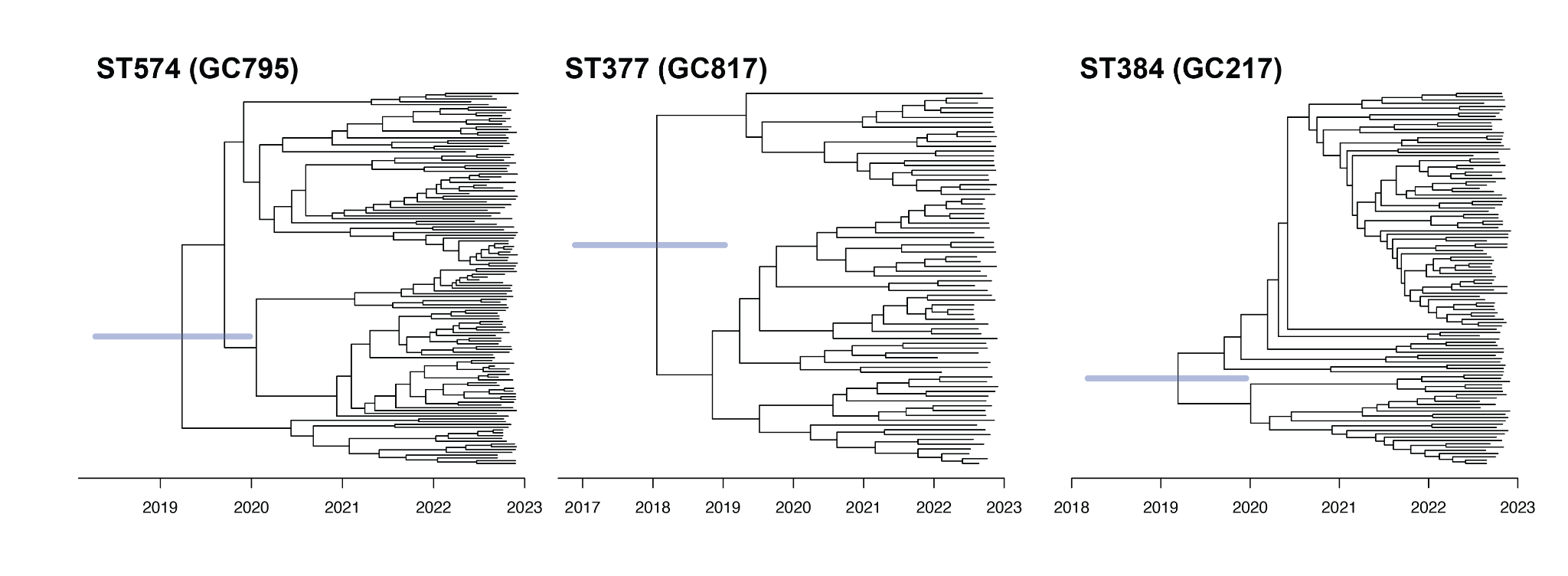

##### **Figure S4. Dated phylogenies for sequence types (ST) and their genomic clusters (GC): ST574 (GC795), ST377 (GC817) and ST384 (GC217).**

The three largest GCs were used to calculate the likely temporal origin of the common ancestors. Dated trees against were inferred using BactDating with the 95% credible intervals illustrated at the root in blue. X-axis: years.

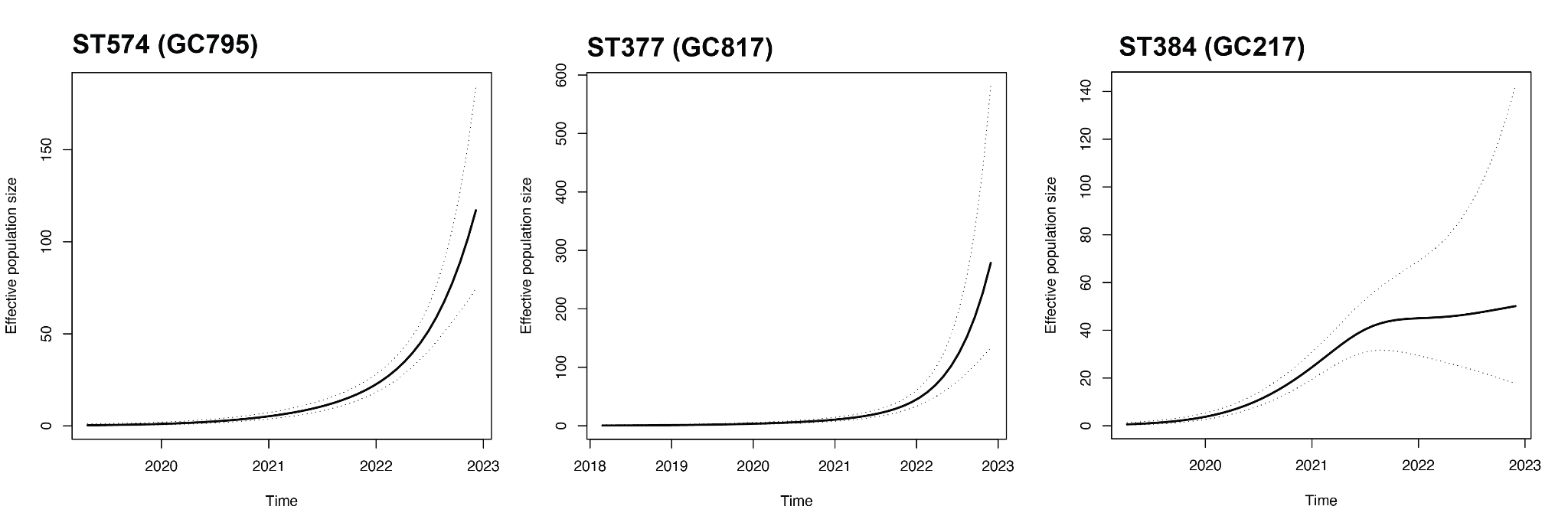

##### **Figure S5. Skygrowth plots of population dynamics of the main outbreak clusters ST574 (GC795), ST377 (GC817) and ST384 (GC217).**

The population size history, calculated with skygrowth, showed significant exponential growth in 2022 for GC795 and GC817. For GC817, the disproportionate increase in population growth started a little earlier (2021) but was not as pronounced as for GC795 and GC817.
